# Reserpine maintains photoreceptor survival in retinal ciliopathy by resolving proteostasis imbalance and ciliogenesis defects

**DOI:** 10.1101/2022.09.14.22279917

**Authors:** Holly Y. Chen, Manju Swaroop, Samantha Papal, Anupam K. Mondal, Gregory J. Tawa, Florian Regent, Hiroko Shimada, Kunio Nagashima, Natalia de Val, Samuel G. Jacobson, Wei Zheng, Anand Swaroop

**Affiliations:** Neurobiology, Neurodegeneration and Repair Laboratory, National Eye Institute, National Institutes of Health; Bethesda, MD 20815, USA; National Therapeutics for Rare and Neglected Diseases, National Center for Advancing Translational Sciences, National Institutes of Health; Rockville, MD 20850, USA; Electron Microscopy Laboratory, National Cancer Institute, Center for Cancer Research, Leidos Biomedical Research, Frederick National Laboratory; Frederick, MD 21701, USA; Department of Ophthalmology, Scheie Eye Institute, Perelman School of Medicine, University of Pennsylvania; Philadelphia, PA 19104, USA

**Author notes:** Department of Cell, Developmental and Integrative Biology, University of Alabama at Birmingham; Birmingham, AL 35233, USA. Department of Physiology, Keio University School of Medicine; Tokyo 160-8582, Japan (Hiroko Shimada). These authors contributed equally (Manju Swaroop, Samantha Papal, Anupam K. Mondal).

## Abstract

Ciliopathies manifest from sensory abnormalities to syndromic disorders with multiorgan pathologies, with retinal degeneration a highly penetrant phenotype. Photoreceptor cell death is a major cause of incurable blindness in retinal ciliopathies. To identify drug candidates to maintain photoreceptor survival, we performed an unbiased, high-throughput screening of over 6,000 bioactive small molecules using retinal organoids differentiated from induced pluripotent stem cells (iPSC) of *rd16* mouse, which is a model of Leber congenital amaurosis (LCA)10 caused by mutations in the cilia-centrosomal gene *CEP290*. We identified five non-toxic positive hits, including the lead molecule reserpine, which improved photoreceptor survival in *rd16* organoids. Reserpine also maintained photoreceptors in retinal organoids derived from induced pluripotent stem cells of *LCA10* patients and in *rd16* mouse retina *in vivo*. Reserpine-treated patient organoids revealed modulation of signaling pathways related to cell survival/death, metabolism, and proteostasis. Further investigation uncovered misregulation of autophagy associated with compromised primary cilium biogenesis in patient organoids and *rd16* mouse retina. Reserpine partially restored the balance between autophagy and the ubiquitin-proteasome system, at least in part by increasing the cargo adaptor p62 and improving primary cilium assembly. Our study identifies effective drug candidates in preclinical studies of *CEP290* retinal ciliopathies through cross-species drug discovery using iPSC-derived organoids, highlights the impact of proteostasis in the pathogenesis of ciliopathies, and provides new insights for treatments of retinal neurodegeneration.

## INTRODUCTION

Once considered vestigial, the primary cilium has emerged as a key microtubule-based organelle that senses external environment and modulates diverse signaling pathways in multiple tissues. Aberrant cilium biogenesis and/or ciliary transport and functions lead to numerous diseases, collectively termed ciliopathies, which manifest from sensory abnormalities to syndromic disorders with multi-organ pathologies including aberrant kidney morphogenesis, brain malformation, and congenital retinal degeneration (*1*). The mammalian retina is an extension of the central nervous system that is specialized for vision (*2*). The visual information is captured by rod and cone photoreceptors, integrated and processed by interneurons, and transmitted to the cortex through the retinal ganglion cells. Inability of the retinal photoreceptors to detect and/or transmit light-triggered signals is a major cause of vision impairment in retinal and macular degenerative diseases (*3–5*). Mutations in over 200 genes can lead to inherited retinal diseases (IRDs) (RetNET, https://sph.uth.edu/retnet/), with a combined prevalence of 1/3-4000 individuals (*5, 6*). Among them, up to 20% of IRD-causing genes are involved in primary cilium biogenesis or functions (*7*). Due to extensive clinical and genetic heterogeneity, high inter- and intrafamily variability, and incomplete penetrance (*8*), treatment options for IRD are limited (*9*). Only one gene therapy drug is currently approved by FDA for Leber congenital amaurosis (LCA, MIM204000) caused by *RPE65* mutations (*10*). While the initial clinical outcomes were promising, the long-term data of this gene therapy drug are less encouraging (*11*). Furthermore, development of individualized gene therapy protocols for divergent mutations in a multitude of genes for rare IRDs would be time-consuming, expensive, and labor-intensive (*12*). Thus, gene-agnostic paradigms are being developed for retinal and macular diseases using model organisms and/or stem cell-based approaches (*13*). Small-molecule or antibody drugs represent a relatively affordable and scalable option (*12*). However, over 90% of drug candidates fail in Phase I clinical trials due to the lack of effective model systems which faithfully recapitulate the pathophysiology of human diseases (*14*). The limited number of cells in the retina and challenges in the maintenance of primary retinal cultures have also hindered the progress of therapeutic development.

LCA is a clinically severe and genetically heterogeneous group of IRDs, leading to vision loss in early childhood (*15*). *LCA10* caused by mutations in the cilia-centrosomal gene *CEP290* (also called *NPHP6*) is one of the most common, accounting for over 20% of patients (*16*). Mutations in *CEP290* exhibit pleiotropy with phenotypes ranging from LCA (affecting vision and other sensory systems) to nephronophthisis, and Joubert and Meckel syndromes involving multiple organ systems (*17–21*). The large 290 kDa centrosome-cilia protein CEP290 is ubiquitously expressed and localized to the Y-links of the transition zone of primary cilia, where it acts as a hub for connecting major protein complexes and likely performs a gating function (*17, 22–24*). The *rd16* mouse carries an in-frame deletion in the myosin tail of CEP290, which causes malformed connecting cilium (equivalent to the transition zone) and rudimentary outer segment (the primary cilium of photoreceptor) structure, leading to rapid degeneration of photoreceptors (*25, 26*). Additional studies have also demonstrated a critical role of CEP290 in cilia biogenesis (*24, 27–30*). Thus, *LCA10* is considered a retinal ciliopathy caused by hypomorphic mutations (*31, 32*) that eliminate some functions of CEP290 (*33*). Vision impairment in early childhood of *CEP290*-LCA patients imposes a considerable burden on families and society (*34*). Notably, these patients demonstrate sparing of the foveal cones even at late stages of life, providing an attractive target for therapy (*35*). Gene replacement using AAV vector is difficult because of the large coding region of *CEP290*. Thus, other strategies including antisense oligonucleotides are currently under investigation (*36–41*). However, no approved treatment is currently available for alleviating vision loss due to CEP290 defects.

Generation of three dimensional tissue organoids from induced pluripotent stem cells (iPSCs) (*42, 43*) has revolutionized biological and disease-modeling investigations and created opportunities for high-throughput screening (HTS) to design novel treatment paradigms (*44*). Further refinements of human retinal organoid culture protocols have permitted high efficiency and yield, developmental staging and higher reproducibility (*45–48*). Biogenesis of diverse cell types in retinal organoid cultures largely recapitulate structural and temporal development of *in vivo* human retina and can display intrinsic light responses mimicking those of primate fovea (*49–51*). Patient iPSC-derived human retinal organoids demonstrate disease-associated phenotypes and are being used to evaluate various therapeutic approaches (*52, 53*). However, long and tedious generation protocols spanning 150+ days, inherent variability, and lack of compatible HTS assays still pose challenges for the application of human retinal organoids for developing therapies (*54*).

In this study, we aimed to establish a reliable drug discovery pipeline using an organoid-based HTS platform with a goal to identify drug candidates for maintaining photoreceptor survival in retinopathies, focusing initially on *LCA10*. For primary HTS of over 6000 bioactive small molecules, we designed survival assays using *rd16* mouse iPSC-derived retinal organoids, which showed compromised photoreceptor development and viability. An FDA-approved small molecule drug reserpine was identified as an efficacious lead compound, which also enhanced both rod and cone photoreceptor development/survival in *LCA10* patient iPSC-derived retinal organoids as well as in the *rd16* retina *in vivo*. Transcriptomic analyses of drug-treated patient organoids indicated modulation of cell survival pathways including p53 and proteostasis by reserpine. Further examination validated mis-regulation of autophagy in patient organoids and *rd16* retina and demonstrated partial restoration of proteostasis and improved ciliogenesis after reserpine treatment. Our study thus establishes a cross-species drug discovery pipeline using organoid-based HTS and identifies a repurposing drug candidate for maintaining photoreceptor survival in *LCA10* patients. As the action mechanisms reserpine is partially through the modulation of proteostasis, which could be a common pathological impact of ciliopathies, reserpine and its derivatives could potentially serve as an effect treatment for patients with other retinal ciliopathies.

## RESULTS

### Establishment of an organoid-based HTS platform

We designed an unbiased HTS assay to identify drug candidates that might improve photoreceptor development and/or survival. For a primary screen, we chose mouse retinal organoids because of a much shorter photoreceptor differentiation time and efficient generation of rudimentary outer segments using our modified HIPRO protocol (*55*). We decided to use an iPSC line derived from the *Nrl*-GFP containing *rd16* mice, which express green fluorescent protein (GFP) in all rods and carry an in-frame deletion in the myosin tail domain of CEP290. The *rd16* mice are a model of photoreceptor degeneration observed in *LCA10* patients (*25*) (fig. S1A). The *rd16* mouse derived-iPSCs displayed similar morphology, proliferation rate, and stem cell properties as those derived from the wild type (WT) (fig. S1B and S1C). WT and *rd16* retinal organoids also showed comparable morphology at early stages of differentiation (Fig. 1A). However, at later stages, rod photoreceptors in *rd16* organoids displayed aberrant morphology, with malformed or missing ciliary axoneme, connecting cilium and ciliary rootlets (fig. S1D). We then performed flow cytometry analyses to evaluate differentiation of GFP+ rod photoreceptors in the *rd16* organoid cultures, with a goal to identify quantifiable phenotypes for HTS. The *rd16* organoids demonstrated as much as 50% lower organoid viability and almost 60% fewer GFP+ rod photoreceptors at day (D) 26 (Fig. 1B), when photoreceptor outer segment biogenesis begins in mouse organoids (*56*).

**Fig. 1.**
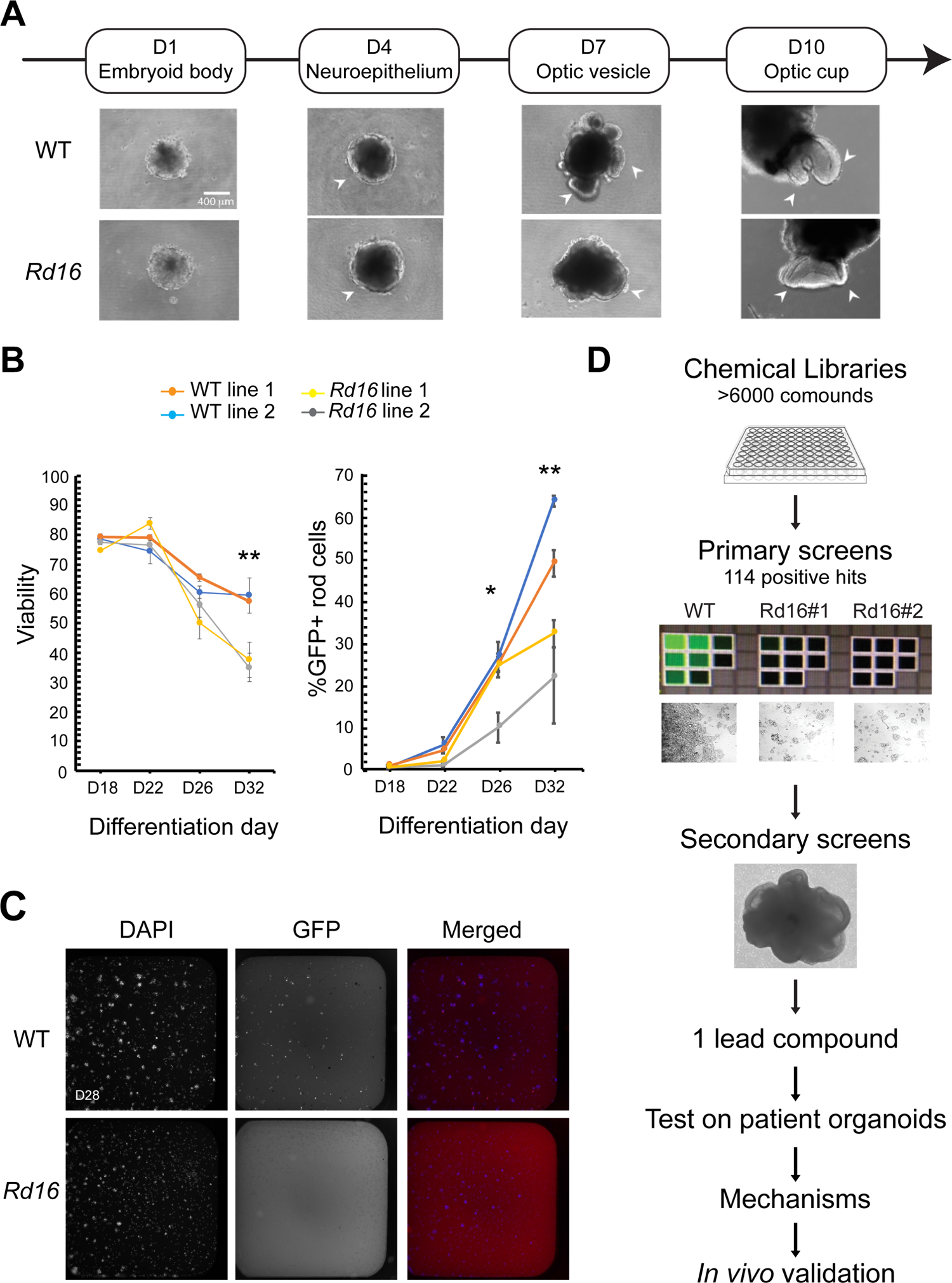
Drug discovery pipelines to identify drug candidates. (**A**) Morphology of *Nrl*-GFP wild type (WT) and *rd16* retinal organoids differentiated from mouse induced pluripotent stem cells (iPSC) at various differentiation time points. (**B**) Flow cytometry analysis of GFP+ rod photoreceptors at different developmental stages. *, p<0.05; **, p<0.01. Each data point summarized at least 3 batches of independent experiments, each of which included at least 10 organoids. (**C**) Fluorescent images of dissociated day (D) 28 WT and rd16 cells stained by 4’,6-diamidino-2-phenylindole (DAPI) and anti-GFP antibody. (**D**) Schematic outline of the drug discovery strategy.

These phenotypes in GFP+ *rd16* rod photoreceptors permitted the development of a HTS screening platform to identify bioactive small-molecule drug candidates for augmenting rod cell differentiation and/or survival (fig. S1E). To avoid variability of organoid cultures, we dissociated *rd16* organoids into single cells at D25 and performed the drug treatment on two-dimensional cultures from D26 to D28. The two-dimensional cultures recapitulated the phenotypes detected in non-dissociated organoids, and *rd16* cells displayed lower viability and fewer GFP+ rod cells at D28 compared to the WT (Fig. 1C).

Our drug discovery pipeline is illustrated in Fig. 1D. In the primary screens, *rd16* retinal organoid-derived cells were treated with over 6,000 small molecules; of these, 114 compounds revealed higher GFP signal intensity, indicating a positive effect on rod photoreceptor survival. We then eliminated the compounds that showed toxicity or autofluorescence even in the absence of GFP expression (false positives). Fourteen small molecule compounds were then selected for further screening to identify a lead compound.

### Selection of the lead compound

We then treated *rd16* retinal organoids with the 14 selected small molecules in a secondary assay, which started at D22 and lasted for 3 days to maximize the drug effect without causing toxicity (Fig. 2A). WT, untreated and treated *rd16* organoids were harvested at D29 to allow sufficient period for restoration of photoreceptors. The lead compound was selected based on the efficacy of the small molecule drugs to improve photoreceptor development and/or survival. Immunostaining of rhodopsin and S-opsin, markers of rod and cone photoreceptors respectively, was performed to quantify the drug effect (Fig. 2B). Rhodopsin is highly expressed and polarized to the apical side of rod photoreceptors in the neural retina of WT organoids. In contrast, *rd16* photoreceptors exhibited lower expression of rhodopsin and the polarity was considerably diminished. Two of the compounds, B05 and to some extent B03, were able to enhance rhodopsin expression, with B05 improving the polarity of expression in neural retina as well. We note that S-cone photoreceptors are difficult to be maintained in mouse organoid culture (*56*); nonetheless, compounds B01 and B05 were able to augment the expression and polarity of S-opsin. Three of the 5 compounds (B01, B03, B05) that showed no toxicity revealed a significant increase in rod and/or cone photoreceptors in D29 *rd16* organoids (Fig. 2C); of these, B05 was the most effective molecule and chosen as the lead compound. Interestingly, the other 4 molecules (B01-B04) were found to be derivatives of B05, further validating our results. B05 was identified as reserpine, a small-molecule drug that has been approved by FDA for treatment of hypertension and schizophrenia (*57, 58*), thereby holding a potential for drug repurposing (Fig. 2D).

**Fig. 2.**
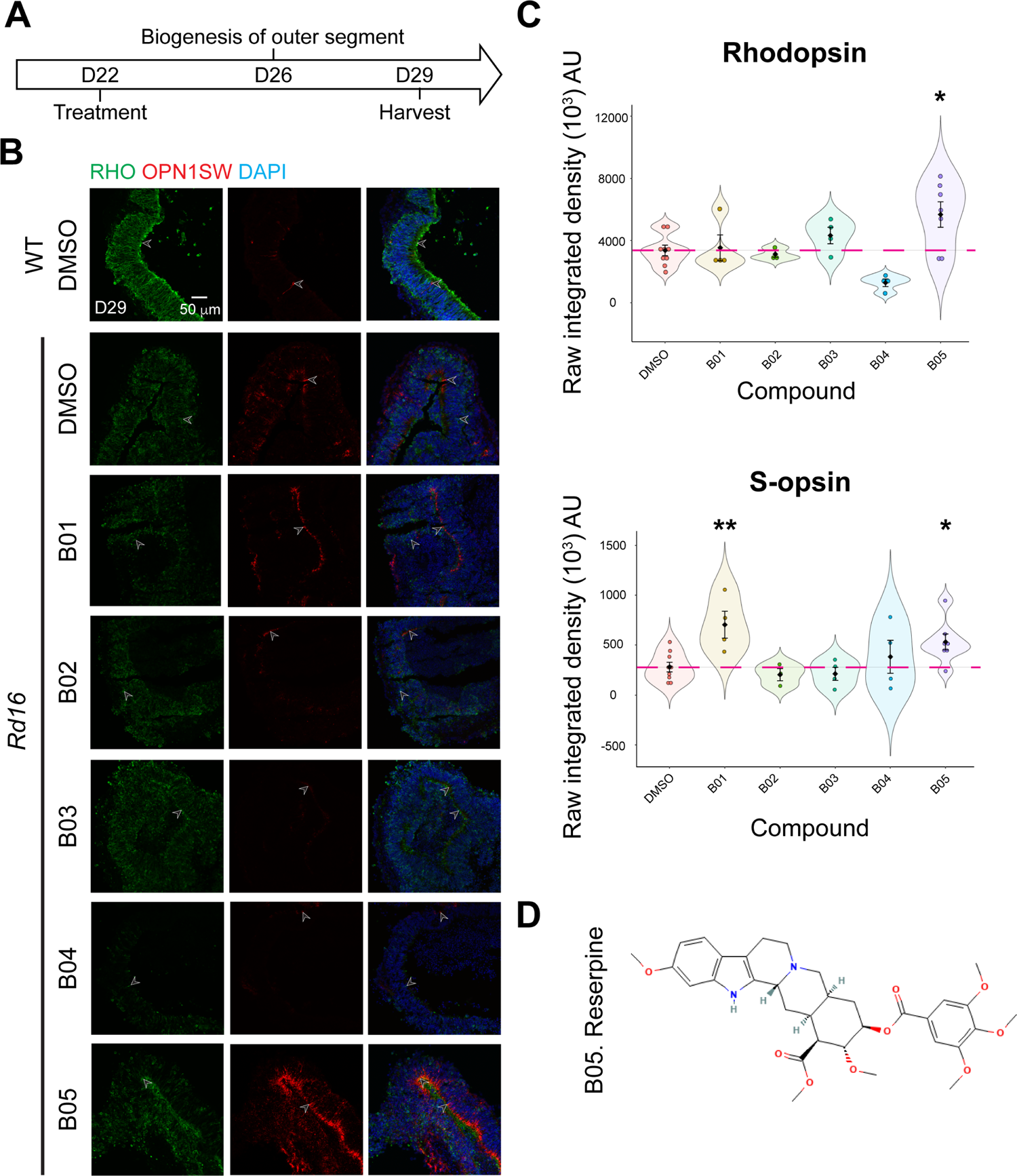
Identification of reserpine as the lead compound. (**A**) Timeline for hit validation in *rd16* retinal organoids. (**B**) Immunostaining of rod cell marker rhodopsin (RHO, green) and cone cell marker S-opsin (OPN1SW, red) in wild-type (WT) and rd16 organoids treated with non-toxic positive hits (B01-B05). Drug vehicle dimethylsulfoxide (DMSO) was used as control. Nuclei were stained by 4’,6-diamidino-2-phenylindole (DAPI). Arrowheads indicate relevant staining. Images were representative of at least 3 independent experiments, each of which had at least 3 organoids. (**C**) Bee swarm plots showing the quantification of fluorescence intensity of rhodopsin (upper) and S-opsin (lower) staining in the validation. The shape of the plot indicates the distribution of data points, which are shown by colorful circles in the center. The black diamond indicates the mean, and the error bar reveals standard deviation. The pink dash line shows the mean fluorescence intensity of DMSO-treated organoids. The plot summarizes at least 3 independent experiments with at least 3 organoids in each batch. *, p<0.05; **, p<0.01. (**D**) Chemical structure depiction of the selected lead compound reserpine.

### Validation of reserpine on *LCA10* patient organoids

To further examine the efficacy of the lead compound reserpine, we utilized iPSC-derived retinal organoids from a previously-reported *LCA10* family (*33*), which comprised of a heterozygous unaffected mother (referred as control henceforth) and two affected compound heterozygous children (LCA-1 and LCA-2; fig. S2A and S2B). Immunoblot analysis of control and patient organoids revealed a significant reduction of full-length CEP290 in patients (fig. S2C). Defects in rod photoreceptor development and outer segment biogenesis were evident in the patient organoids from D120 onwards as revealed by near loss of connecting cilium marker ARL13B and absence or mislocalization of rod-specific protein rhodopsin (fig. S2D). In concordance with the retained central cones in *LCA10*, cone photoreceptors were only slightly compromised at a late stage of differentiation in patient organoids.

Given that photoreceptor outer segment biogenesis begins around D120 in organoids (*47*) and aberrant phenotypes in patient organoids were detectable at this stage (fig. S2D), reserpine was added to patient organoids at D107 at a concentration of 10 μM, 20 μM or 30 μM based on the EC_50_ (half maximal effective concentration) in the primary screens (16.7 μM) (Fig. 3A). We could detect improved connecting cilium immunostaining and rod development as early as D125 with addition of 30 μM reserpine (fig. S3A). Even using the lowest tested dose (10 μM), we observed more polarized rhodopsin and connecting cilium at the apical side of photoreceptors in treated patient organoids, demonstrating a favorable effect of the drug (fig. S3A). The highest tested dose of 30 μM showed no toxicity in most batches of organoid differentiation. We note that reserpine reportedly interferes with the sympathetic nervous system by inhibiting the transport of neurotransmitters into presynaptic vesicles (*59*); yet, even at 30 μM, no adverse effect was detected on the development of ribbon synapses and presynaptic vesicles. In contrast, we observed improved biogenesis of these structures in the outer plexiform layer of treated patient organoids (fig. S3B). Additionally, no significant difference was evident in bipolar cells and Müller glia of untreated and treated patient organoids (fig. S3B). Interestingly, reserpine-treated patient organoids showed reduced GFAP immunostaining (fig. S3B), suggesting reduced cellular stress upon drug treatment. We also noted variations in drug effects on organoids derived from the two patients. While LCA-1 photoreceptors showed more opsin+ rod and cone photoreceptors with the addition of 30 μM reserpine, the improvement in LCA-2 organoids was moderate (Fig. 3B). Such differences could be attributed to variability in response to dose and/or treatment windows for the two donor organoids.

**Fig. 3.**
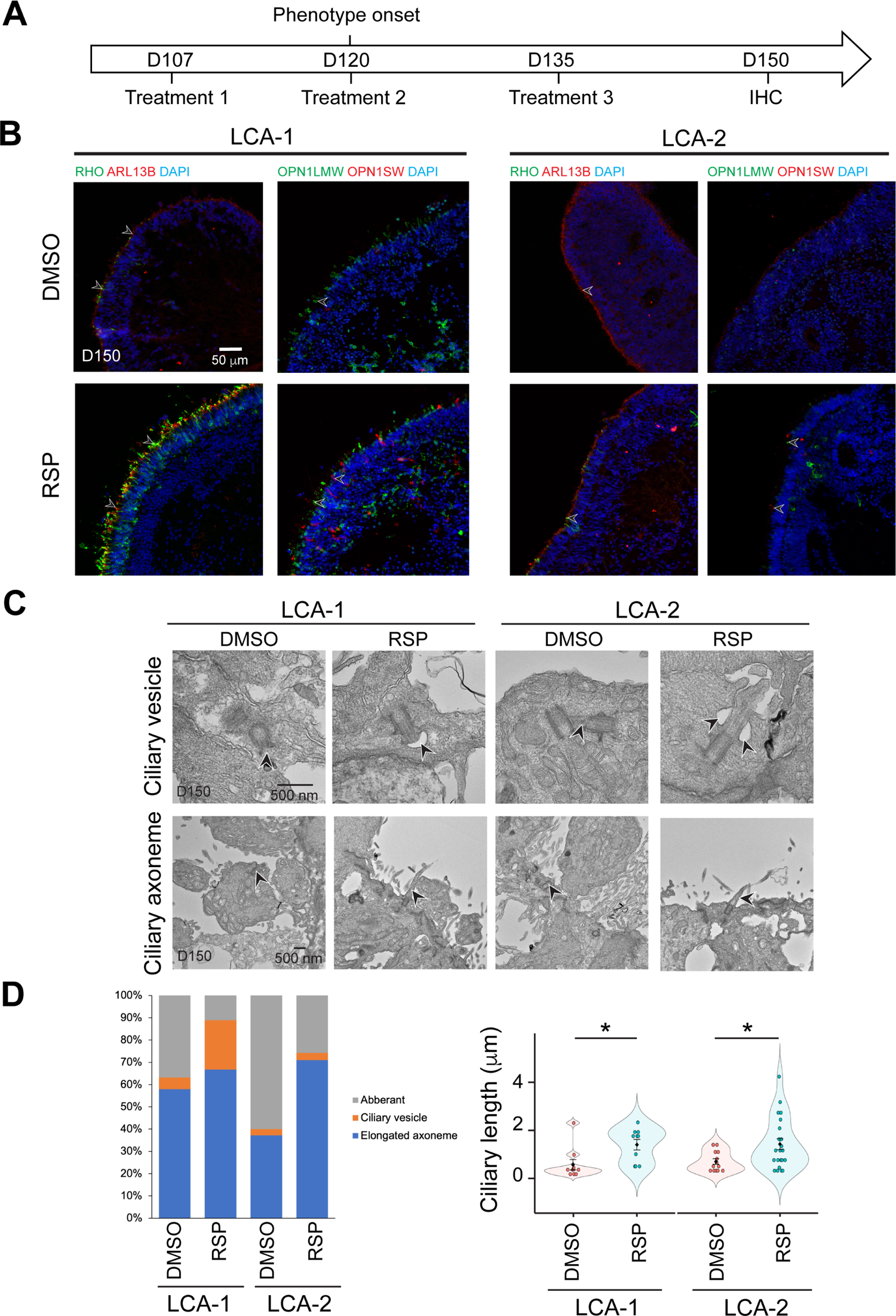
Effect of reserpine (RSP) on *LCA10* patient retinal organoids. (**A**) Timeline for RSP treatments and harvest of patient organoids. (**B**) Immunostaining of rhodopsin (RHO, green) and ARL13B (red) as well as S-opsin (OPN1SW, green) and L/M-opsin (OPN1LMW, red). Nuclei were stained by 4’,6-diamidino-2-phenylindole (DAPI). Arrowheads indicate relevant staining. Images were representative of at least 3 independent experiments, each of which had at least 3 organoids. (**C**) Transmission electron microscopy analysis of control, untreated and treated patient organoids. Arrowheads indicate relevant staining. (**D**) Quantification of the number of the ciliary vesicles, elongated ciliary axoneme and aberrant cilia (left) as well as the length of the primary cilia (right) in untreated and RSP-treated patient organoids. Aberrant cilia were defined as docked mother centrioles without ciliary vesicles or elongated ciliary axoneme. The data summarized at least 4 batches of independent experiments, each of which has at least 2 organoids and 7 docked mother centrioles. The shape of the bee swamp plot indicates the distribution of data points, which are shown by colorful circles in the center. The black diamond indicates the mean, and the error bar reveals standard deviation. *, p<0.05.

We then performed transmission electron microscopy to uncover additional structural details of the primary cilium in photoreceptors of patient organoids after treatment. Reserpine increased the percentage of mother centrioles harboring ciliary vesicles in LCA-1 patient organoids (Fig. 3C and fig. S4A), which were reportedly missing in a significant fraction of LCA photoreceptors (*33*). LCA-2 organoids demonstrated almost 50% more elongated ciliary axonemes after reserpine treatment though ciliary vesicles were hardly noticed. Quantification of the length of the ciliary axoneme indicated a significant increase in both patient organoids after treatment (Fig. 3C). Notably, well-organized disc-like structures could be distinguished in treated samples (fig. S4B), suggesting a positive impact of reserpine treatment on photoreceptor primary cilium development.

### Implication of cell survival and proteostasis pathways in reserpine-treated organoids

To elucidate the mechanism of action of reserpine and to gain insights into photoreceptor cell death in *LCA10*, we performed RNA-seq analyses of control and patient retinal organoids. The organoids were harvested right after the treatment with 30 μM reserpine at D150 (fig. S5A). Consistent with the moderate response of LCA-2 patient organoids to reserpine (Fig. 3B), we did not identify significant differentially expressed (DE) genes between untreated and treated groups of LCA-2 for downstream analyses (data not shown) and thereby focused only on LCA-1. Principal component analysis (PCA) revealed substantial alterations in patient organoid transcriptome compared to control and after reserpine treatment (Fig. 4A). The largest principal component PC1 accounted for up to 36.2% of the total variation and is likely due to the drug treatment. PC2 explained another 21% of the total variation and seemed to be mainly contributed by the disease status. Notably, the reserpine-treated group was closer to the control compared to the untreated group in PC2. A total of 356 genes were significantly differentially expressed between untreated and treated organoids at thresholds of 5% FDR and 2-fold change. Consistent with the PCA plot, heatmap analysis indicated that DE genes in treated patient organoids showed relative transcriptomic shift from untreated ones towards the control ones (Fig. 4B). However, compared to the controls, most of the downregulated genes (e.g., those involved proteostasis) in untreated LCA-1 organoids had an even higher expression upon drug treatment. Similarly, the expression of genes upregulated in untreated samples was lowered even more than the controls by reserpine treatment. Curiously, treatment of LCA-1 organoids with 10 μM or 20 μM reserpine did not yield sufficient DE genes for further analyses (data not shown).

**Fig. 4.**
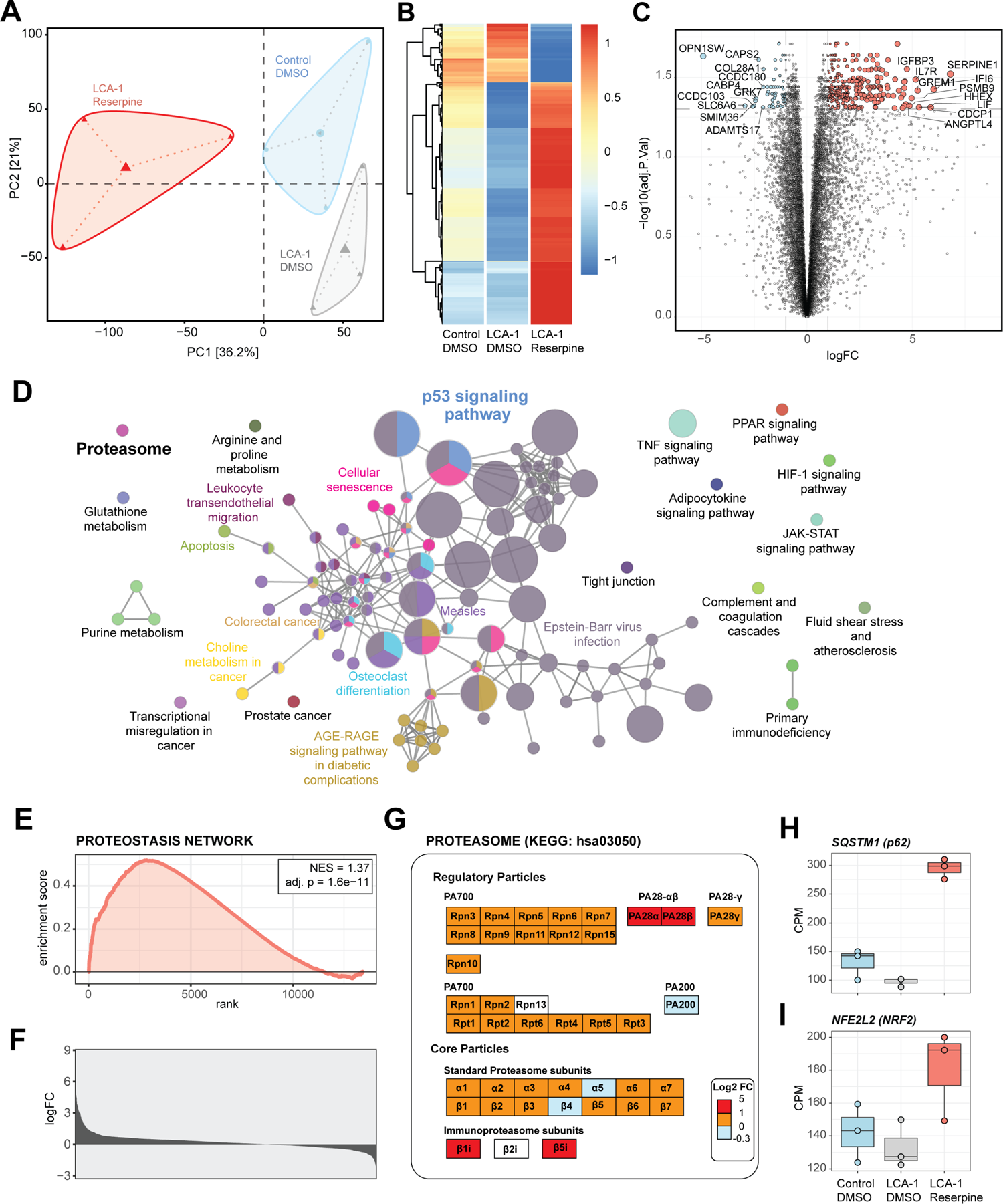
Transcriptomic upregulation of proteasomal components induced by reserpine in patient retinal organoids. (**A**) PCA diagram of patient and control organoids shows altered retinal transcriptomes after reserpine treatment. (**B**) Drug-induced genes in patient organoids displayed specific trends compared to control organoids. (**C**) Volcano plot summarizes reserpine induced differential gene expression changes in LCA-1 organoids. (**D**) ClueGO analysis of KEGG pathway enrichment shows overexpressed genes mapping to protein homeostasis, metabolism and cellular signaling processes. The red rectangle highlights the “Proteasome” factor. (**E**) GSEA plot showing enrichment and significance of Proteostasis Network. (**F**) Histogram of log fold change of the Proteostasis Network genes upon reserpine treatment. (**G**) Proteasomal subunits responding strongly to reserpine treatment. (H) *SQSTM1* (p62) and *NFE2L2* (NRF2), two key regulators of protein homeostasis, showed increased expression after reserpine treatment.

We first analyzed the expression level of genes specific for different retinal cell types and those involved in phototransduction and photoreceptor outer segment structure/function. Reserpine treatment enhanced the expression of marker genes for inner retina cell types, including Müller glia (fig. S5B); however, surprisingly, several rod and cone photoreceptor genes (e.g., *OPN1SW* and *GRK7*) showed lower expression (Fig. 4C, fig. S5C), with *CEP290*-associated cilia genes (*23*) exhibiting varying changes in expression (fig. S5D).

As reserpine did not seem to directly regulate the expression of key components involved in disease pathology, we mapped the overexpressed genes to KEGG pathways and created an enrichment network. Using a cutoff of 3 genes minimum overlap and 5% impact, the annotation network was plotted to visualize leading ontology terms (Fig. 4D). Clusters of metabolism, proteostasis, and immune pathways, together with cell survival related processes, were apparent in the network. To better understand functional connectivity among DE genes, we performed Random-walktrap analysis on their protein-protein interactions network to identify co-functioning over- and under-expressing genes and identified three prominent modules (fig. S5E). Functional module 1 was comprised of extracellular matrix (ECM) and ECM-receptor interaction, and advanced glycation end products (AGE)-Receptor for AGE (RAGE) signaling pathway genes. Müller glia-specific genes *MMPL14*, *TIMP1*, and *VIM* were included in this module. Functional module 2 consisted of inflammation- and proteasome-related genes, which are consistent with the reported role of reserpine as an autophagy modulator (*60*). Notably, functional module 3 contained critical cell survival factors such as p53 signaling and cellular senescence associated genes. We further investigated the trend of the p53 network to characterize its role in response to drug treatment. Expression of *TP53*, the gene coding p53, was found to increase and match the level of control organoids (fig. S5F). In addition, a widespread modulation of each component of the p53 signaling network was evident upon reserpine treatment (fig. S5G). Downstream targets of p53, including metabolic modulator TSC and mTOR complex genes, responded to reserpine and returned to the level of control organoids (fig. S5H). Two components of the mTORC1 signaling pathway, *RHEB* and *LAMTOR1*, were over-expressed after the drug treatment. As mTORC1 is a key regulator of cellular metabolism, we observed significantly elevated expression of *SLC2A1* (GLUT) which is the primary glucose transporter in neurons (fig. S5I).

Given the reported actions of reserpine in neuronal cells (*60*), consistent with activation of mTORC1 activation (*61*) and “proteasome” subunit-encoding genes (Fig. 4D), we performed a focused gene set enrichment analysis to test the impact on the proteostasis network (PN). A PN geneset was manually curated from KEGG and Reactome using keywords reviewed from previous publications (*62, 63*). We detected a significant positive net enrichment of PN in patient organoids after reserpine treatment (NES = 1.37, adj. p-value = 1.6e-11; Fig. 4E), as measured by log_2_ fold change in expression of all PN genes (Fig. 4F). We also identified global overexpression of proteasomal subunits, some of which were notably induced with fold change > 2 (Fig. 4G). In concordance, expression of the key regulators of proteostasis and members of the p53 network, p62 (*SQSTM1*) and NRF2 (*NFE2L2*), were significantly augmented in reserpine-treated patient organoids (Fig. 4H and 4I).

### Restoration of proteostasis in patient photoreceptors

To experimentally examine the role of PN in survival of *LCA10* photoreceptors, we supplemented organoid cultures with various autophagy inhibitors (MRT68921, Lys05, chloroquine, hydroxychloroquine, ROC-325) that target different steps of the autophagy pathway (fig. S6A). MRT68921 and Lys05 inhibit the initiation of autophagy (*64*) and were highly toxic even at one fourth of the reported EC_50_. Curiously, hydroxychloroquine and ROC-325 but not chloroquine enhanced the number of RHO+ cells in patient organoids with more polarized localization of rhodopsin to the apical side (fig. S6B). Small molecule ROC-325, a more specific and efficient autophagy inhibitor compared to hydroxychloroquine (*65*), demonstrated a more dramatic effect on the rescue. Though hydroxychloroquine and ROC-325 were not as efficacious as reserpine (fig. S6B), improvement in rod photoreceptors upon treatment provided further evidence in support of the autophagy pathway as a target for designing therapies.

To further determine the role of autophagy, we evaluated the level of cargo receptor sequestosome 1 (SQTM1) or p62, which recognizes cellular components and helps in formation of autophagosomes for proteolysis (*66*)(Fig. 5A). We noted that LCA-1 patient organoids had a significantly lower level of p62 compared to the control, and reserpine treatment significantly elevated p62 levels by more than 2-fold (Fig. 5B), consistent with previous studies (*60, 67*). Significantly higher levels of LC3-II in patient organoids also indicated accumulation of autophagosomes (defects in autophagy), and reserpine treatment revealed a trend of decreasing LC3-II levels (though not statistically significant) in patient organoids (Fig. 5B). The comparable ratio of LC3-II/LC3-I indicated a similar rate of autophagosome formation between control and patient organoids, suggesting that increased LC3-II levels in patient organoids could be due to overexpression of autophagy pathway components. Consistent with the reported function of reserpine as an autophagy inhibitor, the treated patient organoids had a significantly lower LC3-II/LC3-I ratio, although we did not detect a significant difference between the control and patient organoids (Fig. 5B).

**Fig. 5.**
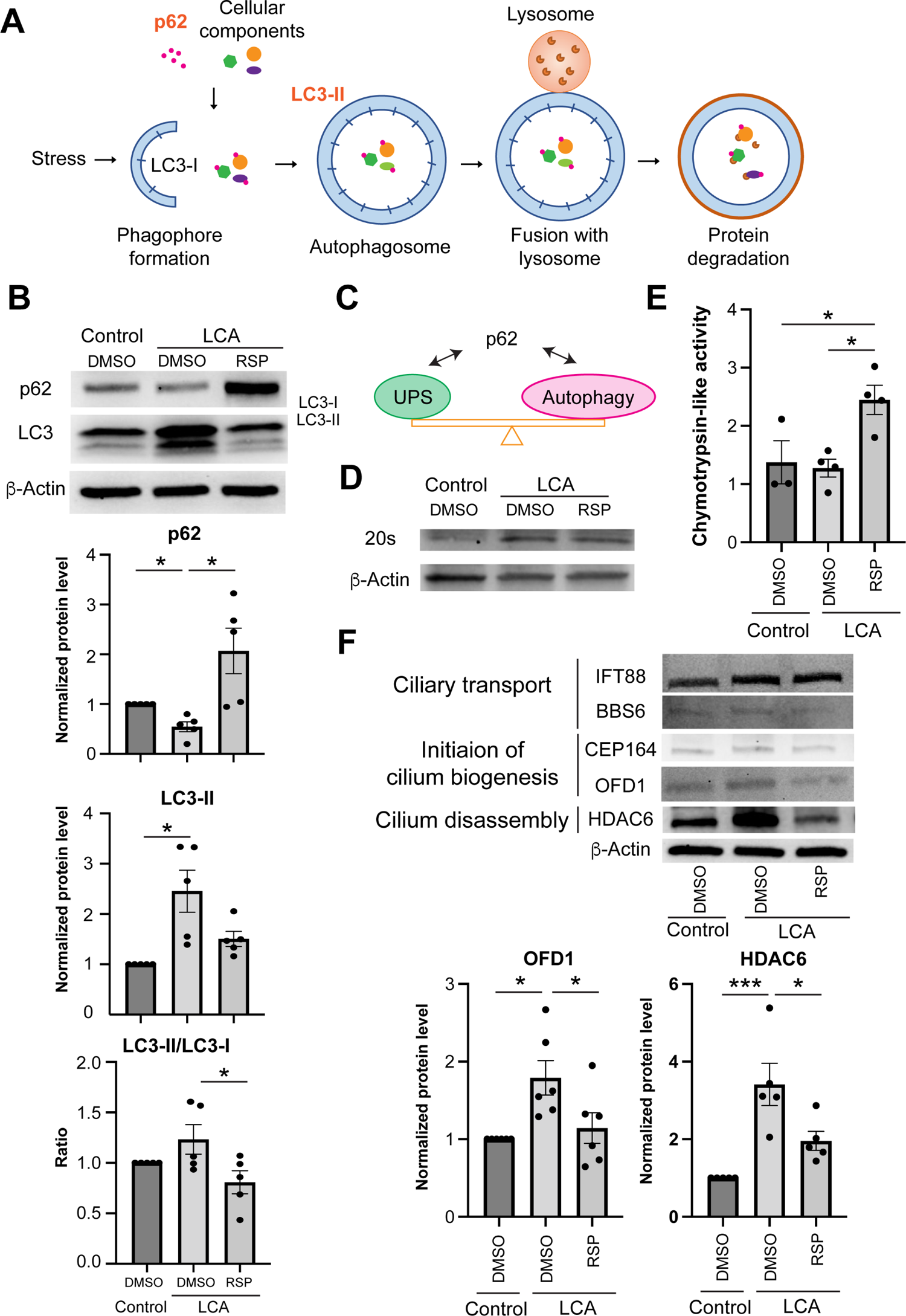
Proteostasis Network in patient organoid cultures in response to reserpine treatment. (**A**) Schematic diagram of autophagy. (**B**) Immunoblot analyses and quantification of autophagy cargo adaptor p62 and autophagosome marker LC-II in control and patient organoids treated with reserpine (RSP). (**C**) Schematic diagram showing the proteome balance between ubiquitin-proteasome system (UPS) and autophagy is mediated through p62 as documented in the literature. (**D**) Immunoblot analysis of the 20S proteasome in control, DMSO-, and RSP-treated cultures. β-Actin was used as the loading control. (**E**) Proteasomal chymotrypsin-like activity in organoids. (**F**) Immunoblot analyses and quantification of key regulators of cilium assembly/disassembly in control, untreated and RSP-treated patient organoids. The drug vehicle dimethylsulfoxide (DMSO) was added to the cultures in the untreated group at the same volume as the drugs. β-Actin was used as the loading control. The histograms summarize data in at least 3 batches of experiments, each of which had at least 3 retinal organoids per group. Each dot in the histogram shows data in one batch of experiment and are presented as mean ± standard deviation. *, *p*<0.05; ***, *p*<0.005.

We then applied Bafilomycin A1 (BafA1), an inhibitor of autophagosome-lysosome fusion, to patient organoid cultures. A short 6-hour treatment with BafA1 did not alter the autophagy pathway in the control, as shown by comparable levels of p62 and LC3-II as well as LC3-II/LC3-I ratio (fig. S6C); however, patient organoids demonstrated a significant (up to 70%) increase of LC3-II and LC3-II/LC3-I ratio, suggesting an elevated autophagic flux in patient organoids, consistent with the rescue by reserpine treatment.

The ubiquitin-proteasome system (UPS) and autophagy are the two key pathways in proteostasis and are reported to interact through p62 (*68, 69*) (Fig. 5C). We therefore investigated the response of UPS in patient organoids upon reserpine treatment. Untreated patient organoids revealed a high level of 20S proteasome, which could be barely detected in the control (Fig. 5D). Nonetheless, both the control and untreated patient organoids showed comparable total catalytic activity and reserpine significantly elevated the proteasome activity (Fig. 5E). These results suggest that increase in 20S expression is likely a compensatory mechanism for compromised proteasome activity in patient organoids, and that reserpine facilitates the clearance of accumulated cellular components and/or autophagosome (Fig. 5B).

We were intrigued by the reported links of autophagy to primary cilium biogenesis (*70–72*) and therefore looked at the expression of key regulators involved in ciliary transport, cilium assembly and disassembly (Fig. 5F). We identified higher expression of OFD1 (oral-facial-digital syndrome 1), which when eliminated by autophagy is shown to promote ciliogenesis (*71*), in patient organoids compared to the control even though autophagy activity was higher in the latter (Fig. 5F). This apparent ambiguity could be due to variations in autophagic adapter machinery for cargo identification. Nonetheless, reserpine treatment reduced OFD1 levels in patient organoids and should facilitate initiation of cilia biogenesis. Another key regulator, histone deacetylase 6 (HDAC6), which deacetylates microtubules and destabilizes the primary cilium for disassembly (*73, 74*), was significantly elevated in patient organoids compared to the control. Dramatic reduction of HDAC6 by reserpine (Fig. 5F) would also have a favorable impact on cilia biogenesis. Consistent with this hypothesis, the addition of a selective HDAC6 inhibitor Tubastatin A to patient organoids improved rod photoreceptor development as shown by a higher number of RHO+ cells (fig. S6D). Tubastatin A treatment enhanced the polarity of not only rhodopsin but also S-opsin, likely due to increased stability of intracellular microtubules and improved intracellular trafficking.

### Maintenance of photoreceptor survival in *rd16* mice *in vivo*

We then performed intravitreal injection of 40 μM reserpine into *rd16* mouse eyes at postnatal day (P)4, and the retinas were harvested at P21 (Fig. 6A). Reserpine was able to maintain photoreceptor survival, with a significantly thicker outer nuclear layer (ONL) in both central and peripheral retina (Fig. 6B). As a small molecule drug, reserpine is highly diffusive, and we observed the effect of drug in contralateral eyes of the treated mice (fig. S7A). Indeed, bilateral therapeutic effects have been reported following unilateral injection in clinical treatments (*75-79*). Notably, no toxicity was observed in the treated mice (data not shown).

**Fig. 6.**
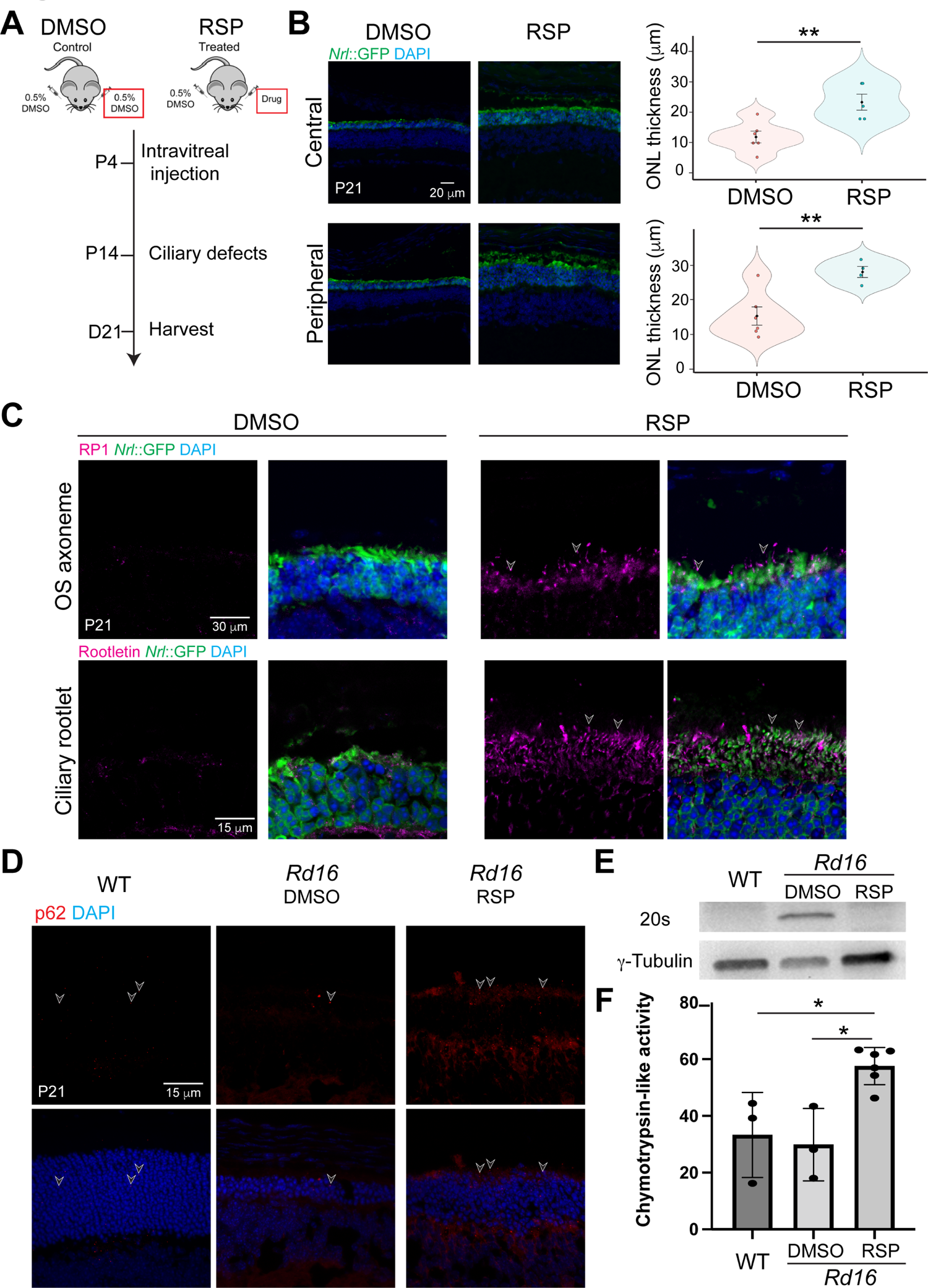
Intravitreal injection of reserpine (RSP) into *rd16* mice. (**A**) Timeline of *in vivo* intravitreal injection. The eyes used for subsequent analyses were highlighted by red rectangles. (**B**) Immunostaining of DMSO- and RSP-treated *rd16* retina (left) and quantification of the GFP+ outer nuclear layer (ONL) thickness (right). The shape of the bee swamp plot indicates the distribution of data points, which are shown by colorful circles in the center. The black diamond indicates the mean, and the error bar reveals standard deviation. *, *p*<0.05. (**C**) Immunostaining of outer segment (OS) axoneme marker RP1 (magenta, upper) and ciliary rootlet marker Rootletin (magenta, lower). (**D**) p62 level in DMSO- and RSP-treated photoreceptors shown by immunostaining. In (**B**), (**C**), and (**D**), nuclei were stained by 4’,6-diamidino-2-phenylindole (DAPI). Arrowheads indicate relevant staining. Images were representative of at least 3 mice from different litters. (**E**) Western blot analysis of 20S proteasome in wild type (WT), DMSO-, and RSP-treated retina of *rd16* mice. γ-Tubulin was used as the loading control. (**F**) Proteasomal chymotrypsin-like activity in *rd16* retina. Data in the histogram were summarized from at least 3 mice from different litters and are presented as mean ± standard deviation. *, *p*<0.05.

Further analyses confirmed an improvement in structure of photoreceptor outer segments of the treated *rd16* retina. Reserpine partially restored outer segment axonemes that were largely missing in untreated mouse retina (Fig. 6C). In addition, substantial ciliary rootlets were conspicuous in the inner segment (shown by GFP) and extended into the ONL. Phototransduction proteins including rhodopsin and Pde6β were transported to the outer segment region (fig. S7B). In addition, the treatment with 40 μM reserpine led to better morphology of other retinal cell types or structures including synapses, bipolar neurons and amacrine cells (fig. S7C). Notably, reserpine treatment significantly reduced GFAP levels that expanded to the entire retina in untreated mice (fig. S7C), suggesting alleviation of photoreceptor stress. Consistent with the effects observed on human retinal organoids, reserpine treatment increased p62 levels (Fig. 6D), dramatically reduced the 20S proteasome (Fig. 6E), and significantly enhanced the proteasome activity (Fig. 6F).

## DISCUSSION

Repurposing of existing drugs for new unrelated clinical modalities provides an excellent opportunity for alleviating patient sufferings in a timely and cost-effective manner (*80*). HTS of approved small molecule drugs can be based on simple assays that target the modulation of a disease-related phenotype or molecule. The use of patient-derived iPSCs and their derivatives, including differentiated cell types in two-dimensional or organoids in three-dimensional cultures, have substantially enhanced the prospects of successful drug discovery (*54, 81*). Exciting screening platforms are now being applied to retinal cells and organoids for drug discovery (*44, 82*). We note the success of cross-species screening, in which the drug candidates identified using one species could be effective in another species (*83*). Given the “orphan” status of IRDs, extensive genetic and phenotypic heterogeneity, and predominantly early dysfunction/death of rods, we designed a simple assay using GFP-tagged rods from iPSCs of a mouse mutant that phenocopies *LCA10* and established an HTS platform to identify small-molecule drugs to maintain photoreceptor survival. As two-dimensional primary or stem cell-derived cultures demonstrate a relatively lower variation and have generated promising candidates in screenings (*44, 84–86*), we dissociated mouse retinal organoids into single cells and performed HTS in two-dimensional cultures. The identified lead compound reserpine was subsequently verified to be effective on patient organoids and in mouse mutant retina *in vivo*, suggesting the feasibility of our approach for drug discovery.

Reserpine was approved for treatment of hypertension in 1955 and later for treatment of schizophrenia (www.pubchem.gov); however, many better tolerated and more potent hypertensive medications have become available during the last few decades. Though potential side effects of inhibition of presynaptic vesicle formation and consequently depression have been described for reserpine treatment (*87*), we did not observe adverse consequences on ribbon synapses or presynaptic vesicles in treated organoids or *in vivo* mouse retina (fig. S3B and Fig. 7). Our results are consistent with a previous study demonstrating little effect of reserpine on central sympathetic activity in cats (*88*). Additionally, the doses of reserpine for treatment of hypertension or schizophrenia in adults range from 8 μg/kg to 80 μg/kg for a 60 kg adult (89), which are higher than the dose we used in mouse studies (9.6 μg/kg). Besides, local delivery by intravitreal injection or via eye drops should be sufficient for reserpine to elicit its effects in the retina due to highly diffusive property of small-molecule drugs. Thus, we suggest that reserpine could be a safe therapeutic approach for treatment of *LCA10* and probably other retinal ciliopathies. Future studies will focus on toxicity evaluation as well as on identifying more potent and less toxic derivatives of reserpine to initial clinical trials.

**Fig. 7.**
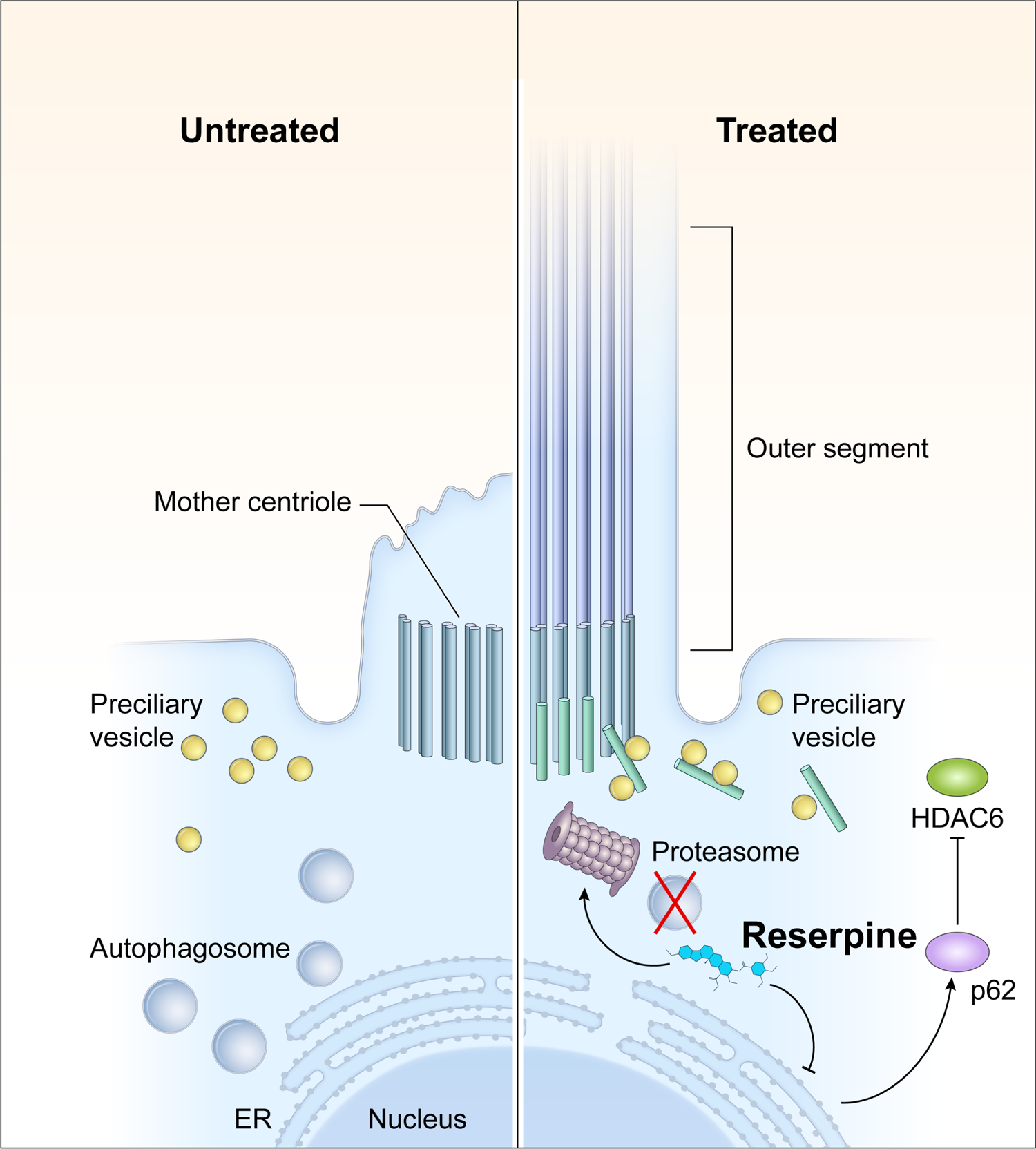
Action mechanisms of reserpine in photoreceptor survival. *CEP29*0 mutations lead to defects in the outer segment biogenesis and consequently ciliary vesicles carrying building blocks of the primary cilium and ciliary proteins are accumulated in patient photoreceptors, leading to the activation of autophagy to degrade unwanted materials. As an autophagy inhibitor, reserpine downregulates autophagy and increases p62 level in photoreceptors. As p62 is a mediator and cargo adaptor of ubiquitin-proteasome system and autophagy, upregulation of p62 not only activates the 20S proteasome to facilitate the clearance of the accumulated autophagosome but also facilitates the degradation of histone deacetylase 6 (HDAC6), which deacetylates microtubules. Removal of HDAC6 in photoreceptors thus should improve the stability of intracellular microtubule and facilitate the transport of pre-ciliary vesicles to the mother centriole for outer segment biogenesis.

Transcriptomic analyses have permitted us to interrogate potential mechanism(s) of reserpine action in patient organoids and implicated signaling pathways involved in immune response (e.g., primary immunodeficiency, complement and coagulation cascade), cell survival and cell death (e.g., p53 signaling pathway, cellular senescence, apoptosis), metabolism (e.g., glutathione metabolism, purine metabolism), and proteostasis. Indeed, these pathways have highly intricate relationships. The p53 protein acts as a sensor of stress conditions and can act both as a transcriptional activator or repressor to promote cell death and cell survival decisions (*90–92*). Gene profiles of treated patient organoids revealed a significant increase of *TP53* as well as its downstream targets including key cellular metabolic regulator TSC and mTOR complex. More importantly, the expression level of genes involved in TSC and mTOR complex returned to the level of control organoids, suggesting a positive impact of reserpine in restoring photoreceptor metabolism. As both TSC knockout and mTOR complex 1 activation are reported to maintain photoreceptor survival (*93, 94*), the trends in our experimental system suggest a cell survival effect of the p53 pathway through the modulation of cell metabolism. In addition, the expression of several Müller glia-specific genes was altered by reserpine, with GFAP levels reduced in both organoids and mouse retina *in vivo*. Müller glia are believed to play a major role in reactive gliosis and likely adapt their transcriptome to support photoreceptors in retinal degeneration (*95, 96*). Yet, we are unsure whether reserpine directly acts on Müller glia to augment photoreceptor survival, or its response is a consequence of improved microenvironment and reduced retinal stress. Notably, the primary cilium appears to have a role in Müller glia maturity and functions in primary cultures (*97*). Whether and how Müller glia are affected in *LCA10* or other degenerative diseases and their role in photoreceptor survival are active areas of investigation.

One of the downstream effectors of p53 and mTOR for metabolic control is the autophagy pathway (*90, 98*). Autophagy is a fundamental homeostatic pathway that serves as a key quality control mechanism for degradation and recycling of components in all tissues, including highly metabolic tissues such as photoreceptors (*99, 100*). Reserpine’s reported role as an autophagy modulator (*60, 101*) is consistent with photoreceptor survival in general (*99, 102–105*), and especially in CEP290 disease because of the purported connection between autophagy and ciliogenesis (*71, 72*). Ciliary defects in photoreceptors promote apoptotic cell death through accumulation of outer segment proteins (e.g., opsin) in the endoplasmic reticulum (ER) and subsequent activation of the unfolded protein response (UPR) (*1*) to restore protein homeostasis in ER. Pharmacological modulation of UPR can preserve photoreceptors in a BBS12 model of syndromic ciliopathy Bardet-Biedl Syndrome (*106*). Recently, CEP290 is shown to be involved in aggresome assembly for protein degradation (*30*), although it is unclear whether this process is disrupted in *LCA10*. Therefore, *CEP290* mutation(s) could lead to accumulation of photoreceptor cilia proteins and ER stress. In agreement, we observed increased autophagic flux in *LCA10* patient organoids. Though it seems counterintuitive to reduce autophagic flux as the accumulated protein would aggravate disease pathology, we hypothesize that reserpine not only acted as an autophagy inhibitor to reduce autophagic flux but also activated UPR to facilitate protein clearance in patient organoids through p62, the common cargo adaptor of the two systems (Fig. 7). We note that inhibition of autophagy or activation of proteasome activity can maintain photoreceptors survival in mouse models with misfolded rhodopsin (*104*). As autophagy defects are associated with retinal degeneration (*99, 105*), reserpine holds promise to serve as a gene agnostic therapy. Interestingly, a favorable effect of autophagy inhibition has also been reported for retinal ganglion cell survival (*107*).

Improved outer segment biogenesis is another prominent effect of reserpine in our study. As a key signaling regulator, the primary cilium is essential for the activation of starvation-induced autophagy through modulation of the Hedgehog pathway and other components involved in autophagy, which can eliminate components in intraflagellar transport (*70*) as well as remove OFD1 from distal appendages to initiate cilium biogenesis (*71*). Thus, the outcome of the crosstalk between ciliogenesis and autophagy seems to largely depend on cell type and physiological context. Our observation of high OFD1 levels in patient organoids compared to the control, despite an elevated autophagic flux, suggests alternative mechanisms. HDAC6 has also been demonstrated to promote autophagosome maturation (*108, 109*) as well as deacetylate microtubules to impede intracellular trafficking, which is a key driver of primary cilium disassembly. Higher expression of HDAC6 in patient organoids could explain both the failure of docking of pre-ciliary vesicles (*33*) and elevated autophagic flux in patient photoreceptors. HDAC6 is shown to be a target of p62 for degradation, and an increase in p62 in photoreceptors can reduce HDAC6 leading to improved outer segment length (*110*). We suggest that reserpine-induced p62 levels can facilitate the degradation of HDAC6 and augment intracellular transport of preciliary vesicles to the mother centriole for cilia biogenesis.

In conclusion, we identified a repurposing small-molecule drug reserpine to maintain photoreceptor survival in retinal ciliopathies, specifically *LCA10*, and at least partially acting by restoration of proteostasis in photoreceptors. Reserpine has been evaluated in a human context (using patient organoids) and mouse retina *in vivo* and thus holds promise in future clinical studies. As the loss of proteomic homeostasis is a major cause of multiple retinal degenerative diseases (*111*), reserpine and its derivatives have clinical potential for gene-agnostic therapies. We realize that we have not examined retinal function and long-term maintenance of photoreceptors in *rd16* mouse. The dose and treatment window of intravitreal injection would require further optimization. In addition, despite the identification of pathways associated with reserpine action, it is hard to dissect the function of each pathway as they are highly intertwined with each other. We should also mention that the drug candidate we identified using mouse organoids is effective on patient organoids and mouse retina *in vivo*; however, certain pathogenetic pathways specific to humans might be missing in the drug target search by current HTS strategy. A major future direction will be to evaluate the effect and safety of reserpine on photoreceptor survival and function in multiple degenerative mouse models and using patient iPSC-derived retinal organoids of other ciliopathies in IRD.

## MATERIALS AND METHODS

### Mouse and human pluripotent cell lines

The mouse *Nrl*-GFP wild-type (WT) and rd16 induced pluripotent stem cell (iPSC) lines were reprogrammed from fibroblasts of day 14.5 mouse embryos as previously described (*55*). The generation and characterization of the familial control, LCA-1 and LCA-2 iPSC lines have been previously reported as well (*33*).

### Animals

*B6.Cg-Cep290rd16/Boc* mice (Strain #: 012283; RRID: IMSR_JAX:012283) were obtained from the Jackson Laboratory and crossed to *Nrlp-EGFP* mice to generate *Nrlp-EGFP*; *Cep290^rd16/rd16^* mice (referred to as *rd16*). The absence of the rd8 mutation in the colony was assessed by PCR. All animal procedures were approved by the Animal Care and Use committee of the National Eye Institutes (Animal study protocol NEI-650) and adhered to ARVO Statement for the Use of Animals in Ophthalmic and Vision Research. Mice were housed in an atmosphere-controlled environment (temperature: 22°C ± 2°C, humidity: 30%–70%), under a 12-hour dark/12-hour light cycle and supplied with food and water ad libitum. Food, water, and nesting material were changed weekly.

### Compound libraries

We used three different small molecule libraries for our screening efforts. The library of 1,280 pharmacologically active compounds (LOPAC 1,280) consists of a collection of small molecules with characterized biological activities commonly used to test and validate screening assays. The library of U.S. Food and Drug Administration (FDA)-2800 approved drugs were set up internally. The compounds were first dissolved in 100% DMSO to generate a stock concentration with a final concentration of 10 mM and subsequently diluted into 7 concentrations and dispensed into 1536-well plates. The NCATS Mechanism Interrogation Plate (MIPE) 5.0 library contains a collection of 1912 compounds (approved for clinical trials).

### Maintenance of induced pluripotent stem cells

The WT and rd16 iPSC lines were maintained as previously described (112). Briefly, the iPSC lines were maintained on feeder cells (Millipore) in maintenance medium constituted by Knockout DMEM (ThermoFisher Scientific), 1x MEM non-essential amino acids (NEAA) (Sigma), 1x GlutaMAX (ThermoFisher Scientific), 1x Penicillin-Streptomycin (PS) (ThermoFisher Scientific), 2000 U/ml leukemia inhibitory factor (LIF) (Millipore), and 15% ES cell-qualified fetal bovine serum (FBS) (ThermoFisher Scientific) at 37°C, 5% CO_2_. Full media change was performed every day, with 55 μM β-Mercaptoethanol (2-ME) (ThermoFisher Scientific) freshly added. Cells were passaged using TrypLE Express (ThermoFisher Scientific) every two days.

Human iPSC lines were maintained in Essential 8 (E8) (ThermoFisher Scientific) on growth factor-reduced (GFR) Matrigel (Corning)-coated plates, with media fully changed daily. Cells were maintained at 37°C, 5% O2, 5% CO_2_ and passaged at 60-80% confluency using the EDTA-based dissociation method (113).

### Differentiation of mouse and human retinal organoids

The modified HIPRO protocol was used to differentiate mouse iPSCs into retinal organoids (55, 112). At differentiation day (D)0, iPSCs were plated in low adhesion U-shaped 96-well plate (Wako) at a density of 3000-5000 cells per well in 100 μl retinal differentiation medium consisting of GMEM (ThermoFisher Scientific), 1x NEAA, 1x sodium pyruvate (Sigma) and 1.5%(v/v) knockout serum replacement (KSR) (ThermoFisher Scientific). At D1, 240 μl GFR-Matrigel (>9.5 mg/ml) was diluted in 1.8 ml retinal differentiation medium and 20 μl diluent was aliquoted to each well of the 96-well plate. Retinal organoids from one 96-well plate were transferred to a 100 mm poly(2-hydroxyethyl methacrylate) (Sigma)-coated petri dish with 12 ml DMEM/F12 with GlutaMAX, 1x N2 supplement and 1x PS. The media were replaced by DMEM/F12 with GlutaMAX, 1x PS, 1x N2 supplement, 1 mM taurine (Sigma), 500 nM 9-*cis* retinal (Sigma) and 100 ng/ml insulin-like growth factor 1 (IGF1) (ThermoFisher Scientific) at D10, and half-media change was performed every other day, with 55 μM 2-ME freshly added to the media. From D26 and onward, 1x NEAA, 1x B27 supplement without Vitamin A (ThermoFisher Scientific) and 2%(v/v) FBS (ThermoFisher Scientific) were added to the culture. Half-media exchanges were performed every two days, with 55 μM 2-ME freshly added to media. The cultures were incubated in 5% O_2_ from D0 to D10 and in 20% O_2_ from D10 onwards.

Human retinal organoid differentiation was performed as previously described (114). Briefly, small clumps dissociated from iPSCs in one well of a 6-well plate were resuspended in E8 medium supplemented with 10 µM Y-27632 (Tocris) and transferred into one 100-mm polyHEMA-coated petri dish for embryoid body (EB) formation. Media were supplied with neural induction media (NIM) (DMEM/F-12 (1:1) (ThermoFisher Scientific), 1x N2 supplement, 1x NEAA, 2 µg/ml heparin (Sigma) at D1 and D2 at a ratio of 3:1 and 1:1 respectively, and fully switched to NIM at D3. D7 EBs from one 100-mm petri dish were plated onto one GFR Matrigel-coated 60 mm dish and cultured in NIM, with media changed every 2-3 days. In the application requiring a large-scale production of retinal organoids, nicotinamide was added to the culture to reach a final concentration of 5 mM from D0 to D8 (*48*). NIM was replaced by 3:1 retinal induction medium (RIM) consisting of DMEM/F-12 supplemented with 1x B27 without Vitamin A (ThermoFisher Scientific), 1% antibiotic-antimycotic solution (ThermoFisher Scientific), 1% GlutaMAX and 1X NEAA at D16 and media change was performed every day until D28, on which the adherent cells were scraped off into small clumps (<5 mm^2^) and split into two polyHEMA-coated 100-mm petri dishes. The floating clumps were cultured in RIM supplemented with IGF1 (ThermoFisher Scientific) and 1 mM taurine (Sigma), with 55 μM 2-ME freshly added. From D38 and onward, 10% fetal bovine serum was added to RIM supplemented with 20 ng/ml IGF-1, and 1 mM taurine. 1 μM 9-*cis* retinal was freshly supplemented to the cultures during media change from D63 to D91. From D91 till the end of differentiation, the concentration of 9-*cis* retinal was reduced to 0.5 μM and B27 without Vitamin A was replaced by N2. Once scraped, media were half changed every 2-3 days, with IGF1, taurine and 9-*cis* retinal freshly added to the media under dim light environment.

### Dissociation of mouse retinal organoids into single cells

Mouse retinal organoids were transferred into a 15 ml centrifuge tube using wide-bored transfer pipets and washed one time with 10 ml 1x PBS (ThermoFisher Scientific). Prewarmed 1 ml 0.25% trypsin-EDTA was then added, and the tube was incubated s at 37°C for 10 minutes, with pipetting up and down for 10 times using P1000 pipetman at 5- and 10-minute incubation. 10 ml retinal maturation media (DMEM/F12 with GlutaMAX, 1x PS, 1x N2 supplement, 1 mM taurine, 500 nM 9-*cis* retinal, 1x NEAA, 1x B27 supplement without Vitamin A, 2%(v/v) FBS) was added to the tube. The tube was inverted several times before centrifugation at 200g for 5 minutes. After removal of supernatant, the cell pellet was resuspended in 1 ml retinal maturation media and filtered through 40-μm cell strainer (BD Bioscience) before proceeding to subsequent analyses.

### Immunoblot analysis

At least 3 organoids in each batch were homogenized in 100 μl of RIPA buffer (Sigma) supplemented with 1x protease inhibitors (Roche) and 1x phosphatase inhibitor (Roche). The lysate was agitated at 4°C for half an hour, before centrifugation at 12,000*g* for 10 minutes at 4°C. The supernatant was either stored at −80 °C until use or quantified by Pierce bicinchoninic acid (BCA) protein assay (ThermoFisher Scientific). Approximately 20 μg protein was diluted 4:1 in reducing 4x Laemmeli buffer (Biorad) and boiled for 10 minutes. The samples were separated at 150-180 V for 1 hour on 4-15% precast polyacrylamide gel (Biorad) and transferred to polyvinylidene fluoride (PVDF) membranes using a TransBlot® Turbo™ Transfer System (Biorad). After blocking in 5% milk or 5% bovine serum albumin (BSA) for 1 hour at room temperature, the blots were incubated in antibody cocktails (Table S1) overnight in 1% milk or BSA in 1X TBS-T at 4 °C overnight with gentle agitation. Membranes were subsequently washed in 1X TBS-T for 3 times, 10 minutes each, and incubated in 1X TBS-T with horseradish peroxidase-conjugated secondary antibodies (1:5,000) for 1 hour at room temperature, followed by another three 10-minute wash. Before imaging, the membranes were exposed to SuperSignal® West Pico enhanced chemiluminescence (ECL) solution (ThermoFisher Scientific) for 5 minutes, and chemiluminescence was captured using a Bio-Rad ChemiDoc™ touch (Bio-Rad).

### Chymotrypsin-like proteasome activity assay

The chymotrypsin-like protease activity associated with the proteasome complex in organoids or mouse retina was quantified using the Proteasome 20S Activity Assay Kit (Sigma) following manufacturer’s protocol. In short, at least 3 retinal organoids or 1 mouse retina were homogenized in 55 μl PBS mixed with 55 μl reconstituted Proteasome Assay Loading Solution and incubated on ice for 30 minutes. After centrifuging at 10,000 *g* for 10 minutes at 4 °C, 10 μl supernatant was taken for BCA assay to determine the protein amount and the remaining 100 μl was transferred to one well of black/clear 96-well plate for incubation at 37 °C. Protease activity of individual samples was measured by the fluorescence intensity (λex = 480–500 nm/λem = 520–530 nm) normalized to the protein amount.

### Transmission electron microscopy

Retinal organoids were processed for transmission electron microscopy (TEM) analysis as previously described (*33*). Briefly, the organoids were fixed in 4% formaldehyde and 2% glutaraldehyde in 0.1 M cacodylate buffer, pH 7.4 (Tousimis) for 2 hours at room temperature, followed by 3 washes in cacodylate buffer before further fixation in osmium tetroxide (1% v/v in 0.1 M cacodylate buffer; Electron Microscopy Sciences) for 1 hour at room temperature. The organoids were then washed in the same buffer for three times, followed by 1 wash in acetate buffer (0.1 M, pH 4.2), and *en-bloc* staining in uranyl acetate (0.5% w/v; Electron Microscopy Sciences) in acetate buffer for 1 hour at room temperature. The samples were dehydrated in a series of ethanol solutions (35%, 50%, 75%, 95%, and 100%) and then by propylene oxide. The samples were subsequently infiltrated in a mixture of propylene oxide and epoxy resin (1:1, v/v) overnight, embedded in a flat mold with pure epoxy resin, and cured at 55 °C for 48 hours. 70-80 nm sections were made with an ultramicrotome (UC 7) and diamond knife (Diatome), attached on a 200-mesh copper grid, and counter-stained in aqueous solution of uranyl acetate (0.5% w/v) and then lead citrate solutions. The thin sections were stabilized by carbon evaporation before the EM examination. The digital images were taken using a digital camera equipped with an electron microscope (H7650) (AMT).

### Flow cytometry

After dissociating retinal organoids, the cells were resuspended in DPBS (ThermoFisher Scientific) containing 1 mM EDTA (Millipore) and filtered through a 40-μm cell strainer. 4’,6-diamidino-2-phenylindole (DAPI) (ThermoFisher Scientific) was added to the samples before being analyzed by FACSAriaII (BD Bioscience). Cell viability was evaluated by integrity (gated by DAPI), size (gated by forward scatter, FSC) and granularity (gated by side scatter, SSC). WT retinal organoids without GFP marker on the same differentiation day were used to set the gating for GFP+ cells.

### RNA extraction and library preparation

Total RNA was purified from homogenized retinal organoids using TriPure isolation reagent (Roche). Quality of isolated RNAs was assessed using Bioanalyzer RNA 6000 nano assays (Agilent) and high-quality total RNA (RNA integrity number ≥7.5) were used for construction of mRNA sequencing library. 100 ng total RNA were used to construct the strand-specific libraries using TruSeq RNA Sample Prep Kit-v2 (Illumina).

### RNA-seq data analysis

RNA sequencing was performed as described (115). Paired-end 125-bp reads were generated using Illumina sequencing. Reads were quality checked and mapped to the reference transcriptome using Kallisto. Alignments were imported into R using tximport for downstream analyses. The edgeR and limma pipeline were employed for differential expression analysis, while pathway annotation was performed using ClueGO (116). Protein interaction data was obtained from STRING (v11.5) with evidence cut off of 700, via the R package STRINGdb (117). Network analysis was performed using the *igraph* package [https://igraph.org] and genes were mapped to pathways using gProfileR (118). Proteostasis network genes were manually collected by pooling KEGG and Reactome pathways with keywords from (*62*) and (*63*), and the proteosome map was adapted from KEGG (hsa03050). Gene set enrichment analysis was done using the R package fgsea. All other plotting and analyses, unless otherwise mentioned, were performed using tidyverse and base R packages.

### Immunohistochemistry

Mouse and human organoids were fixed in 4% PFA (Electron Microscopy Sciences) for 1 hour at room temperature, washed 1 time and cryoprotected in 15% sucrose for at least 2 hours at room temperature, followed by 30% sucrose at 4°C overnight. The next day, organoids were embedded in Shandon M-1 Embedding Matrix (ThermoFisher Scientific). The blocks were sectioned at 10 μm thickness and incubated at room temperature for at least 1 hour, before immunostaining or storage at −80 °C. After incubating in blocking solution (5% donkey serum in PBS) for 1 hour at room temperature, the sections were supplied with diluted primary antibodies (Table S1) at 4°C overnight. After three 10-min washes in PBS, Species-specific secondary antibodies conjugated with Alexa Fluor 488, 568 or 637, together with DAPI, were diluted in blocking solution (1:500; ThermoFisher Scientific) and incubated with the sections for 1 hour at room temperature. After washing in PBS for three times, 10 minutes each, the sections were mounted for imaging.

For mouse retinal sections, rd16 mice of both sexes were injected at postnatal (P) day 4 and recovered at P21 after euthanasia using CO_2_ atmosphere. Eyes were denucleated before being pierced in the center of the cornea using a 26-gauge needle. Eyecups were then incubated for 15 min in 4% paraformaldehyde (PFA). The cornea and lens were then dissected, and the eyecups were placed in 4% PFA for 15 min at room temperature (RT) before proceeding to cryoprotection in 20% and 30% sucrose-PBS at 4C for 1 hour and overnight, respectively. Eyecups were then quickly frozen in Shandon™ M-1 Embedding Matrix (Thermo Fisher Scientific) and cut at 12μm. Retinal sections were washed twice in PBS then blocked in PBS containing 5% Donkey serum and 0.3% Triton X-100 (PBST) for 1 hour at room temperature. Slides were incubated overnight at 4°C with primary antibody diluted in PBST at an appropriate concentration (Table S1). Sections were then washed 3 times with PBS and incubated with secondary antibody and 1 μg/ml 4,6-diamidino-2-phenylindole (DAPI) for 1 hour at room temperature. After 3 washes in PBS, the sections were mounted in Fluoromount-G (SouthernBiotech).

### Image acquisition and analysis

Brightfield images were taken using an EVOS XL Core Cell Imaging System (ThermoFisher). Fluorescence images were acquired with LSM-880 confocal microscope (Zeiss) with Zen software. FIJI and Photoshop CC 2019 software were used for images export, analyzing and processing. Rhodopsin and S-opsin fluorescent intensity of organoid sections were quantified with FIJI using the maximum intensity projections of z-stack images. Multi-channel RGB (red, green, blue) images were separated into 8-bit grayscale images and regions of interest were identified by applying the “Moment” threshold algorithm of FIJI. The same threshold algorithm was used for all images. Area, raw and integrated fluorescence intensity in each image were then quantified with Fiji and plotted using RStudio version 1.1.463.

### High-throughput screening (HTS) assays

To optimize the assay for HTS, D25 WT and *rd16* mouse retinal organoids were dissociated into single-cell suspension and plated in 5 µl retinal maturation medium II at a concentration of 3000, 4000, and 5000 cells per well in a 1536-well plate. After culturing in a 37°C humidified incubator overnight, 5000 cells/well yielded an optimal plating density for both WT and *rd16* cultures and were selected for subsequent assays.

In the screening experiments for drug candidates, dissociated D25 WT-positive control and *rd16* mouse retinal organoids were filtered through 35-μm cell strainer and dispensed into 1536-well plate at a density of 5000 cells/well in 5 µl freshly prepared retinal maturation medium II using the Multidrop Combi Dispenser (Thermo Scientific). Cells were incubated in a 37°C humidified incubator overnight for cell recovery and attachment to the plates. Compounds in the 1536-well drug source plates were added to the 1536-well assay plates at a volume of 23 nl/well using an NX-TR pintool station (WAKO Scientific Solutions). Cells were treated with compounds for 48 hours, followed by addition of 0.5 µl quencher, and GFP and DAPI signal intensities were quantified using the acumen® Cellista (TTP, Labtech). The positive hits with a significant increase of GFP compared to the DMSO-treated cells were retested using dissociated retinal organoids from iPSCs without GFP marker to remove autofluorescent false positives. The remaining drug candidates were further tested using a full 11-concentration setting (1:3 serial dilutions starting at 10 mM) in triplicate plates to prioritize the hits based on efficacy.

The primary and confirmatory screening data were analyzed using software developed internally at the NIH Chemical Genomics Center (NCGC) (119). The plate data was processed as follows: We first ran quality control on the plate data by visual inspection, masking wells showing erroneous signals, e.g., localized groupings of wells exhibiting enhanced or inhibited signal. We then performed intra-plate normalization as follows:

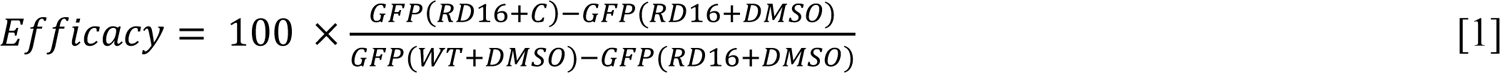

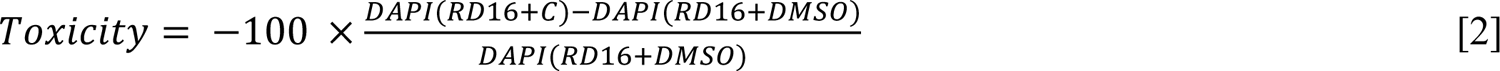

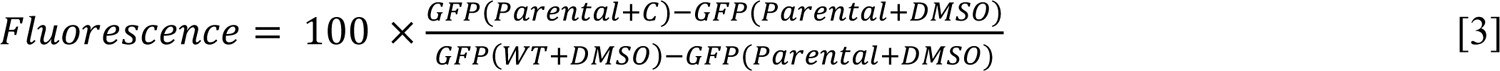

Where:

WT = D25 WT retinal organoid cells with GFP marker = GFP + RD16 = D25 *rd16* retinal organoid cells with GFP marker= GFP - Parental = Retinal organoids from iPSCs without GFP marker = GFP-C = compound tested GFP = GFP (channel 488) signal intensity DAPI = DAPI (channel 405) signal intensity

When the data is normalized in this fashion, efficacy, toxicity, and fluorescence range from 0 to 100%. With this normalization completed we compute dose response curves as follows:

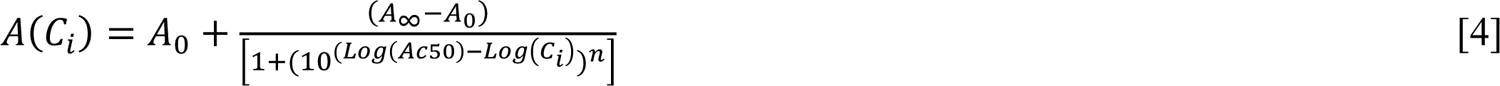

Where *C_i_* = the *i’th* concentration, *A(C_i_)* = activity (efficacy, toxicity, or fluorescence) at concentration *i*, *A_0_* = activity at zero concentration, *A_∞_* = activity at infinite concentration, and *EC_50_* = the concentration giving a response half-way between the fitted top (100%) and bottom (0) of the curve.

Compounds are desired that have high efficacy (*EC_50efficacy_*= small, *A_∞, efficacy=_* high), low

toxicity, (*EC_50toxicity_* = large) and low fluorescence (*EC_50fluorescence_* = large)

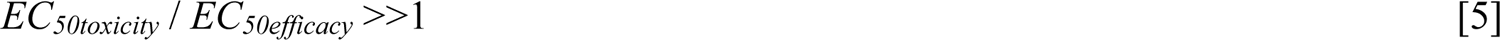

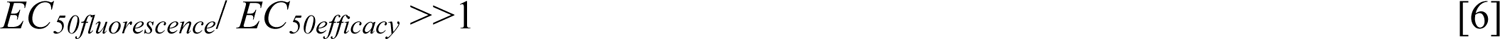

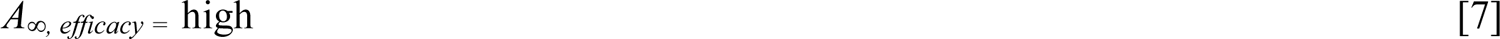

In addition, the dose response curves for each measured quantity must be well behaved, i.e., (i) curves exhibit values near zero at low concentration, (ii) values increase with increasing concentration, (ii) curves show a well-defined inflection point at the *EC_50_* concentration, and (iv) curves show a well-defined plateau at high concentration (119).

### Intravitreal injection

Animal experiments were conducted in the animal facility at National Eye Institute, National Institutes of Health. The facility approved the animal care and experimental procedures used in this study (NEI-ASP650). P4 *Nrl*-GFP *rd16* mice pups of either sex were anesthetized on ice and their eyelids were opened using a 30-gauge needle to gently expose the eye. 1mg/ml Ketoprofen was injected for analgesia. Half microliter of solution diluted in PBS was blindly delivered using a glass micropipette (World Precision Instruments) produced using a Flaming-Brown micropipette puller (Sutter Instruments) and connected to an Eppendorf Femtojet air compressor (Eppendorf). Mice with eye/lens damage following the injection were excluded from further analyses.

### Statistics

Based on the initial data, the sample size of animals used in each group was determined by unpaired t-test to be less than 6 mice in each group (http://www.biomath.info/). For experiments involved mouse or human retinal organoids, at least four independent experiments, each of which had at least 3 organoids, were performed unless specified. For animal studies, at least 2 retina from 1 mouse of at least 4 different litters was evaluated. All data were expressed as mean ± standard deviation (SD) unless specified. An unpaired two-tailed Student’s *t* test was used to compare the mean between two groups. For comparison of three groups, one-way AVOVA followed by Tukey’s test was performed. Results with a *p*-value < 0.05 were considered statistically significant.

## Data Availability

All data needed to evaluate the conclusions in the paper are present in the paper and/or the Supplementary Materials. RNA-seq data are available through GEO accession #206959.

## ACKNOWLEDGMENTS

We thank Tiansen Li, Linn Gieser, Matthew Brooks, Ryan A. Kelley and Trupti Shetty in Neurobiology, Neurodegeneration & Repair Laboratory of the National Eye Institute and Wenwei Huang in National Center for Advancing Translational Sciences for technical support and insightful discussions. We are grateful to Rafael Villasmil, Julie Laux, Jessica Albrecht, and Jacqueline Minehart (Flow Cytometry Core Facility of the National Eye Institute), Lijin Dong, Pinghu Liu, and Jingqi Lei (Genetic Engineering Core Facility of the National Eye Institute), Jizhong Zou and Jeanette Beers (iPSC Core Facility at National Heart, Lung, and Blood Institute), Sandra Burkett (Molecular Cytogenetic Core Facility at National Cancer Institute), and Alan Hoofring and Ethan Tyler (Division of Medical Arts at National Institutes of Health) for assistance in various aspects of research. These studies were supported by the high-performance computational capabilities of the Biowulf Linux cluster at the National Institutes of Health (http://biowulf.nih.gov). The work was supported by National Eye Institute Intramural Research Programs (ZIAEY000450 and ZIAEY000546) and National Center for Advancing Translational Sciences Intramural Research Programs ZIATR000018-06.

## MATERIAL AVAILABILITY STATEMENT

All pluripotent stem cell lines and animals are available upon request. A Material Transfer Agreement complying with the guidelines of National Institutes of Health has to be established before shipment.

## Source Data Files - Legends

**Fig. 5. Source data 1. Overlay of the brightfield and chemiluminescence images indicating the signal of p62. RSP stands for reserpine.** The size of the protein ladders, p62, and relevant sample identity are labeled.

**Fig. 5. Source data 2. Overlay of the brightfield and chemiluminescence images indicating the signal of LC3. RSP stands for reserpine.** The size of the protein ladders, LC3, and relevant sample identity are labeled.

**Fig. 5. Source data 3. Overlay of the brightfield and chemiluminescence images indicating the signal of β-Actin. RSP stands for reserpine.** The size of the protein ladders, β-Actin, and relevant sample identity are labeled.

**Fig. 5. Source data 4. Overlay of the brightfield and chemiluminescence images indicating the signal of 20s proteosome and β-Actin.** RSP stands for reserpine. The size of the protein ladders, β-Actin, 20s proteosome, and relevant sample identity are labeled.

**Fig. 5. Source data 5. Overlay of the brightfield and chemiluminescence images indicating the signal of IFT88. RSP stands for reserpine.** The size of the protein ladders, IFT88, and relevant sample identity are labeled.

**Fig. 5. Source data 6. Overlay of the brightfield and chemiluminescence images indicating the signal of BBS6. RSP stands for reserpine.** The size of the protein ladders, BBS6, and relevant sample identity are labeled.

**Fig. 5. Source data 7. Overlay of the brightfield and chemiluminescence images indicating the signal of CEP164. RSP stands for reserpine.** The size of the protein ladders, CEP164, and relevant sample identity are labeled.

**Fig. 5. Source data 8. Overlay of the brightfield and chemiluminescence images indicating the signal of OFD1. RSP stands for reserpine.** The size of the protein ladders, OFD1, and relevant sample identity are labeled.

**Fig. 5. Source data 9. Overlay of the brightfield and chemiluminescence images indicating the signal of HDAC6. RSP stands for reserpine.** The size of the protein ladders, HDAC6, and relevant sample identity are labeled.

**Fig. 5. Source data 10. Overlay of the brightfield and chemiluminescence images indicating the signal of β-Actin. RSP stands for reserpine.** The size of the protein ladders, β-Actin, and relevant sample identity are labeled.

**Fig. 6. Source data 1. Overlay of the brightfield and chemiluminescence images indicating the signal of 20s proteosome.** WT and RSP stand for wild type and reserpine respectively. The size of the protein ladders, 20s proteosome, and relevant sample identity are labeled.

**Fig. 6. Source data 2. Overlay of the brightfield and chemiluminescence images indicating the signal of γ-Tubulin.** WT and RSP stand for wild type and reserpine respectively. The size of the protein ladders, γ-Tubulin, and relevant sample identity are labeled.

**Fig. S2. Source data 1. Overlay of the brightfield and chemiluminescence images indicating the signal of CEP290.** The size of the protein ladders, CEP290, and relevant sample identity are labeled.

**Fig. S2. Source data 2. Overlay of the brightfield and chemiluminescence images indicating the signal of GAPDH.** The size of the protein ladders, GAPGH, and relevant sample identity are labeled.

**Fig. S5. Source data 1. Overlay of the brightfield and chemiluminescence images indicating the signal of LC3.** The size of the protein ladders, LC3, and relevant sample identity are labeled.

**Fig. S5. Source data 2. Overlay of the brightfield and chemiluminescence images indicating the signal of p62.** The size of the protein ladders, p62, and relevant sample identity are labeled.

**Fig. S5. Source data 3. Overlay of the brightfield and chemiluminescence images indicating the signal of β-actin.** The size of the protein ladders, β-Actin, and relevant sample identity are labeled.

## Supplementary Materials and Methods

### Animals

*Cep290^rd16/rd16^* mice were obtained from the Jackson Labs and crossed to *Nrlp-EGFP* mice to generate *Nrlp-EGFP*; *Cep290^rd16/rd16^* mice (referred to as *rd16*). The absence of the rd8 mutation in the colony was assessed by PCR. All animal procedures were approved by the Animal Care and Use committee of the National Eye Institutes (Animal study protocol NEI-650) and adhered to ARVO Statement for the Use of Animals in Ophthalmic and Vision Research. Mice were housed in an atmosphere-controlled environment (temperature: 22°C ± 2°C, humidity: 30%– 70%), under a 12-hour dark/12-hour light cycle and supplied with food and water ad libitum. Food, water, and nesting material were changed weekly.

### Maintenance of induced pluripotent stem cells

The WT and rd16 iPSC lines were maintained as previously described (*113*). Briefly, the iPSC lines were maintained on feeder cells (Millipore) in maintenance medium constituted by Knockout DMEM (ThermoFisher Scientific), 1x MEM non-essential amino acids (NEAA) (Sigma), 1x GlutaMAX (ThermoFisher Scientific), 1x Penicillin-Streptomycin (PS) (ThermoFisher Scientific), 2000 U/ml leukemia inhibitory factor (LIF) (Millipore), and 15% ES cell-qualified fetal bovine serum (FBS) (ThermoFisher Scientific) at 37°C, 5% CO_2_. Full media change was performed every day, with 55 μM β-Mercaptoethanol (2-ME) (ThermoFisher Scientific) freshly added. Cells were passaged using TrypLE Express (ThermoFisher Scientific) every two days.

Human iPSC lines were maintained in Essential 8 (E8) (ThermoFisher Scientific) on growth factor-reduced (GFR) Matrigel (Corning)-coated plates, with media fully changed daily. Cells were maintained at 37°C, 5% O2, 5% CO_2_ and passaged at 60-80% confluency using the EDTA-based dissociation method (*114*).

### Differentiation of mouse and human retinal organoids

The modified HIPRO protocol was used to differentiate mouse iPSCs into retinal organoids (*55, 113*). At differentiation day (D)0, iPSCs were plated in low adhesion U-shaped 96-well plate (Wako) at a density of 3000-5000 cells per well in 100 μl retinal differentiation medium consisting of GMEM (ThermoFisher Scientific), 1x NEAA, 1x sodium pyruvate (Sigma) and 1.5%(v/v) knockout serum replacement (KSR) (ThermoFisher Scientific). At D1, 240 μl GFR-Matrigel (>9.5 mg/ml) was diluted in 1.8 ml retinal differentiation medium and 20 μl diluent was aliquoted to each well of the 96-well plate. Retinal organoids from one 96-well plate were transferred to a 100 mm poly(2-hydroxyethyl methacrylate) (Sigma)-coated petri dish with 12 ml DMEM/F12 with GlutaMAX, 1x N2 supplement and 1x PS. The media were replaced by DMEM/F12 with GlutaMAX, 1x PS, 1x N2 supplement, 1 mM taurine (Sigma), 500 nM 9-*cis* retinal (Sigma) and 100 ng/ml insulin-like growth factor 1 (IGF1) (ThermoFisher Scientific) at D10, and half-media change was performed every other day, with 55 μM 2-ME freshly added to the media. From D26 and onward, 1x NEAA, 1x B27 supplement without Vitamin A (ThermoFisher Scientific) and 2%(v/v) FBS (ThermoFisher Scientific) were added to the culture. Half-media exchanges were performed every two days, with 55 μM 2-ME freshly added to media. The cultures were incubated in 5% O_2_ from D0 to D10 and in 20% O_2_ from D10 onwards.

Human retinal organoid differentiation was performed as previously described (*115*). Briefly, small clumps dissociated from iPSCs in one well of a 6-well plate were resuspended in E8 medium supplemented with 10 µM Y-27632 (Tocris) and transferred into one 100-mm polyHEMA-coated petri dish for embryoid body (EB) formation. Media were supplied with neural induction media (NIM) (DMEM/F-12 (1:1) (ThermoFisher Scientific), 1x N2 supplement, 1x NEAA, 2 µg/ml heparin (Sigma) at D1 and D2 at a ratio of 3:1 and 1:1 respectively, and fully switched to NIM at D3. D7 EBs from one 100-mm petri dish were plated onto one GFR Matrigel-coated 60 mm dish and cultured in NIM, with media changed every 2-3 days. In the application requiring a large-scale production of retinal organoids, nicotinamide was added to the culture to reach a final concentration of 5 mM from D0 to D8 (*48*). NIM was replaced by 3:1 retinal induction medium (RIM) consisting of DMEM/F-12 supplemented with 1x B27 without Vitamin A (ThermoFisher Scientific), 1% antibiotic-antimycotic solution (ThermoFisher Scientific), 1% GlutaMAX and 1X NEAA at D16 and media change was performed every day until D28, on which the adherent cells were scraped off into small clumps (<5 mm^2^) and split into two polyHEMA-coated 100-mm petri dishes. The floating clumps were cultured in RIM supplemented with IGF1 (ThermoFisher Scientific) and 1 mM taurine (Sigma), with 55 μM 2-ME freshly added. From D38 and onward, 10% fetal bovine serum was added to RIM supplemented with 20 ng/ml IGF-1, and 1 mM taurine. 1 μM 9-*cis* retinal was freshly supplemented to the cultures during media change from D63 to D91. From D91 till the end of differentiation, the concentration of 9-*cis* retinal was reduced to 0.5 μM and B27 without Vitamin A was replaced by N2. Once scraped, media were half changed every 2-3 days, with IGF1, taurine and 9-*cis* retinal freshly added to the media under dim light environment.

### RNA-seq data analysis

RNA sequencing was performed as described (*116*). Paired-end 125-bp reads were generated using Illumina sequencing. Reads were quality checked and mapped to the reference transcriptome using Kallisto. Alignments were imported into R using tximport for downstream analyses. The edgeR and limma pipeline were employed for differential expression analysis, while pathway annotation was performed using ClueGO (*117*). Protein interaction data was obtained from STRING (v11.5) with evidence cut off of 700, via the R package STRINGdb (*118*). Network analysis was performed using the *igraph* package [https://igraph.org] and genes were mapped to pathways using gProfileR (*119*). Proteostasis network genes were manually collected by pooling KEGG and Reactome pathways with keywords from (*62*) and (*63*), and the proteosome map was adapted from KEGG (hsa03050). Gene set enrichment analysis was done using the R package fgsea. All other plotting and analyses, unless otherwise mentioned, were performed using tidyverse and base R packages.

### Immunohistochemistry

For mouse retinal sections, *rd16 mice* of both sexes were injected at postnatal (P) day 4 and recovered at P21 after euthanasia using CO_2_ atmosphere. Eyes were denucleated before being pierced in the center of the cornea using a 26-gauge needle. Eyecups were then incubated for 15 min in 4% paraformaldehyde (PFA). The cornea and lens were then dissected, and the eyecups were placed in 4% PFA for 15 min at room temperature (RT) before proceeding to cryoprotection in 20% and 30% sucrose-PBS at 4C for 1 hour and overnight, respectively. Eyecups were then quickly frozen in Shandon™ M-1 Embedding Matrix (Thermo Fisher Scientific) and cut at 12μm. Retinal sections were washed twice in PBS then blocked in PBS containing 5% Donkey serum and 0.3% Triton X-100 (PBST) for 1 hour at room temperature. Slides were incubated overnight at 4°C with primary antibody diluted in PBST at an appropriate concentration (Table S1). Sections were then washed 3 times with PBS and incubated with secondary antibody and 1 μg/ml 4,6-diamidino-2-phenylindole (DAPI) for 1 hour at room temperature. After 3 washes in PBS, the sections were mounted in Fluoromount-G (SouthernBiotech).

### Image acquisition and analysis

**Fig. S1.**
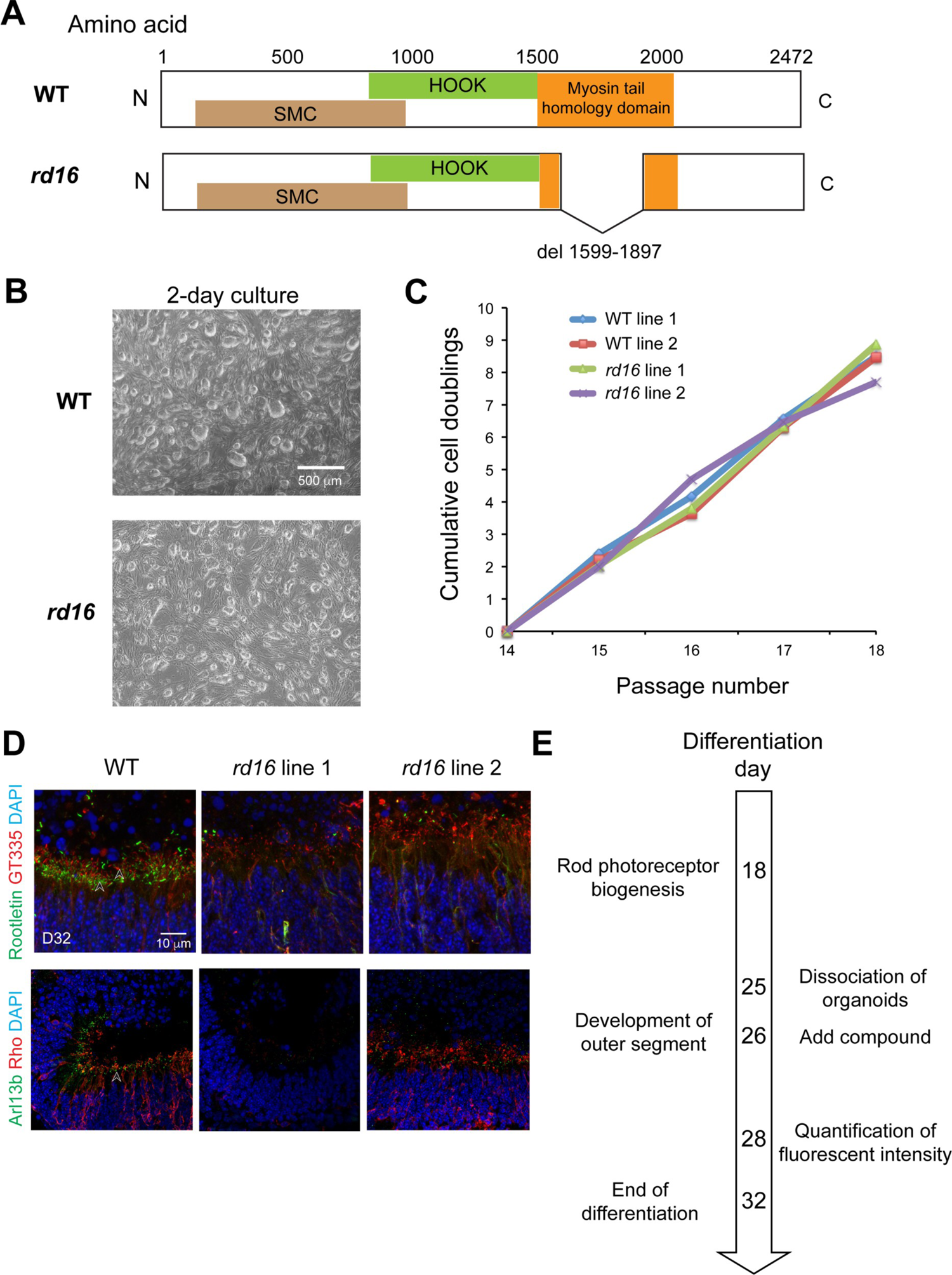
Phenotypes of retinal organoids differentiated from induced pluripotent stem cells of *Nrl*-GFP *rd16* mice. (**A**) Deletion of myosin tail domain in *rd16* mice compared to the wild type (WT) ones. (**B**) Morphology of iPSC colonies, (**C**) proliferation rate of iPSCs, and (**D**) photoreceptor primary cilium of organoids were compared between WT and *rd16*. (**E**) Timeline of the key differentiation events in mouse organoids and primary screens. Images were representative of at least 3 independent experiments, each of which had at least 3 organoids.

**Fig. S2.**
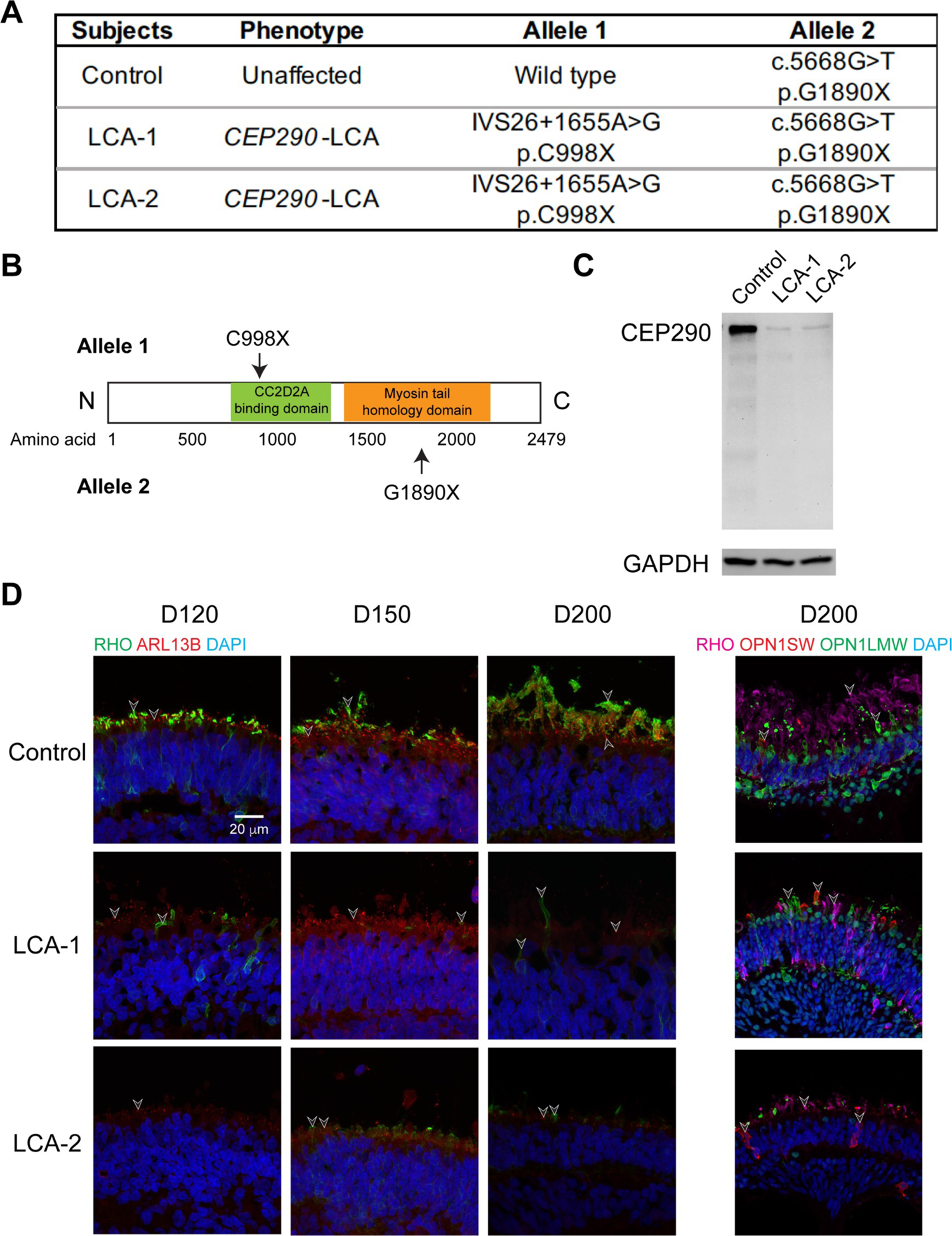
Genotype and phenotype of the *LCA10* family in this study. (**A**) Genotypes of the subjects including a heterozygous but phenotypically normal control and two *LCA10* patients. (**B**) Location of the stop codons caused by point mutations in *LCA10* patients. (**C**) Western blot analysis of CEP290 in day (D) 200 control and patient retinal organoids. Images were representative of at least 3 independent experiments, each of which had at least 3 organoids. GAPDH was used as a loading control. (**D**) Immunostaining of rhodopsin (green) and ARL13B (red) in the left panel as well as rhodopsin (magenta), S-opsin (red) and L/M-opsin (green) in the right panel. Nuclei were stained by 4’,6-diamidino-2-phenylindole (DAPI). Arrowheads indicate relevant staining. Images were representative of at least 3 independent experiments, each of which had at least 3 organoids.

**Fig. S3.**
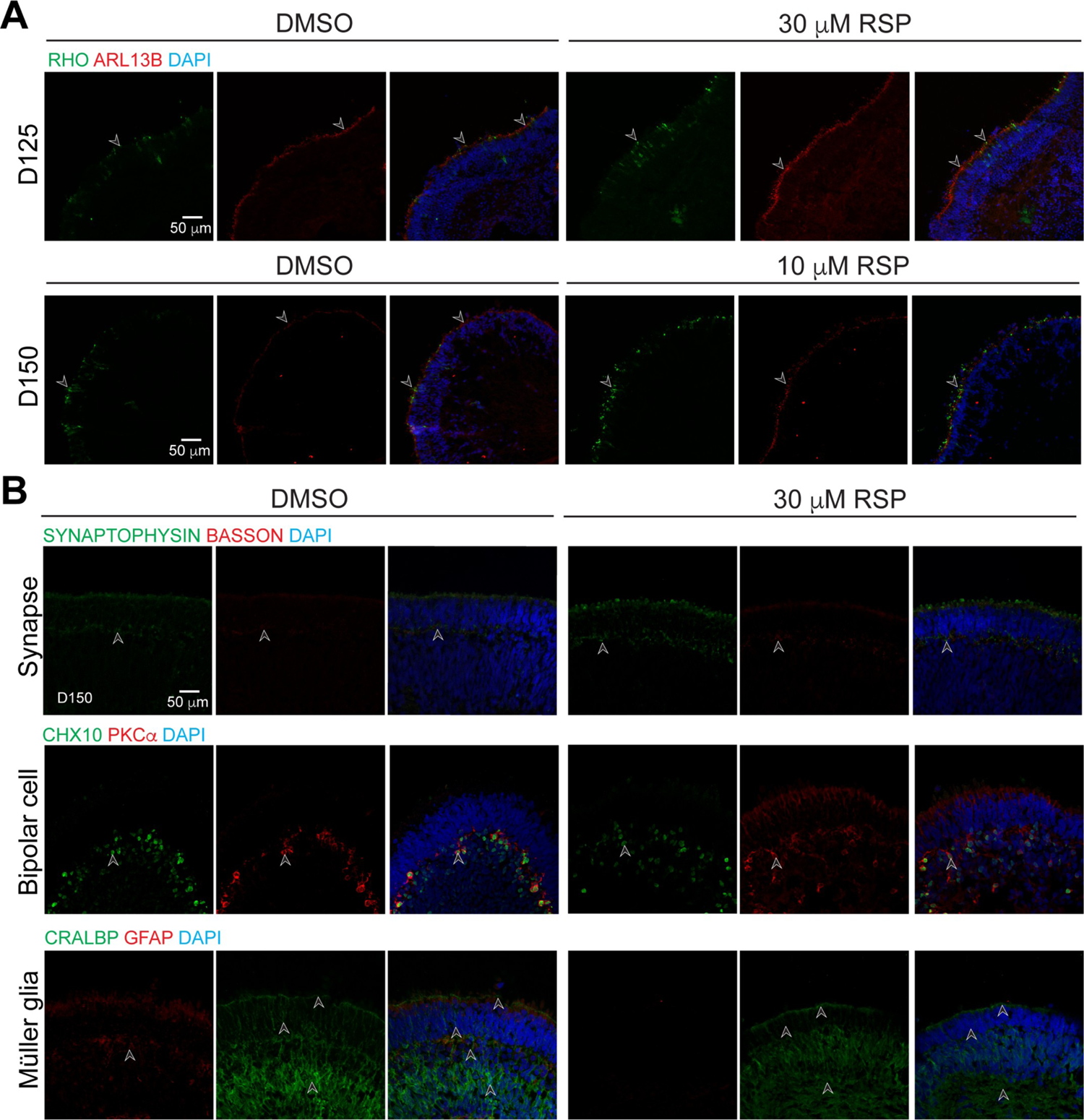
Immunostaining of reserpine (RSP)-treated LCA-1 organoids. (**A**) Rod cell marker rhodopsin (RHO, green) and ciliary marker ARL13B (red) were shown in LCA1 organoids treated with 30 µM RSP at differentiation day (D) 125 (upper) and with 10 µM RSP at D150 (lower). (**B**) Immunostaining of structural or cell type-specific markers including those for ribbon synapses (BASSON, red), presynaptic vesicles (SYNAPTOPHYSIN, green), bipolar cells (CHX10, green; PKCa, red), Müller glia (CRALBP, green; GFAP, red). Nuclei were stained by 4’,6-diamidino-2-phenylindole (DAPI). Arrowheads indicate relevant staining. Images were representative of at least 3 independent experiments, each of which had at least 3 organoids.

**Fig. S4.**
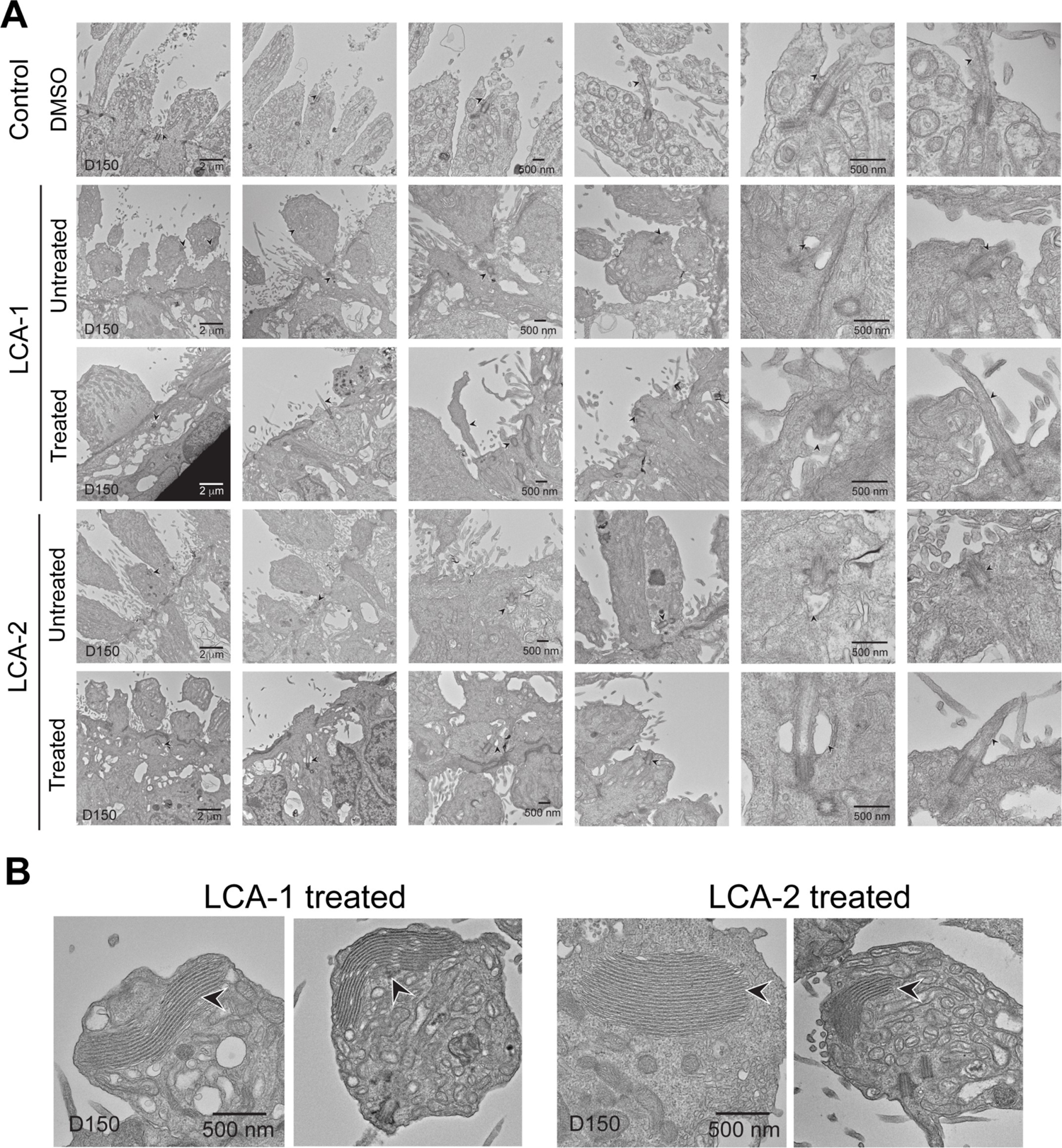
Transmission electron microscopy analyses of control, untreated and treated patient organoids. (**A**) Morphology of photoreceptor primary cilium. (**B**) Stacks of disc-like structures observed in treated patient organoids. Arrowheads indicate relevant structures.

**Fig. S5.**
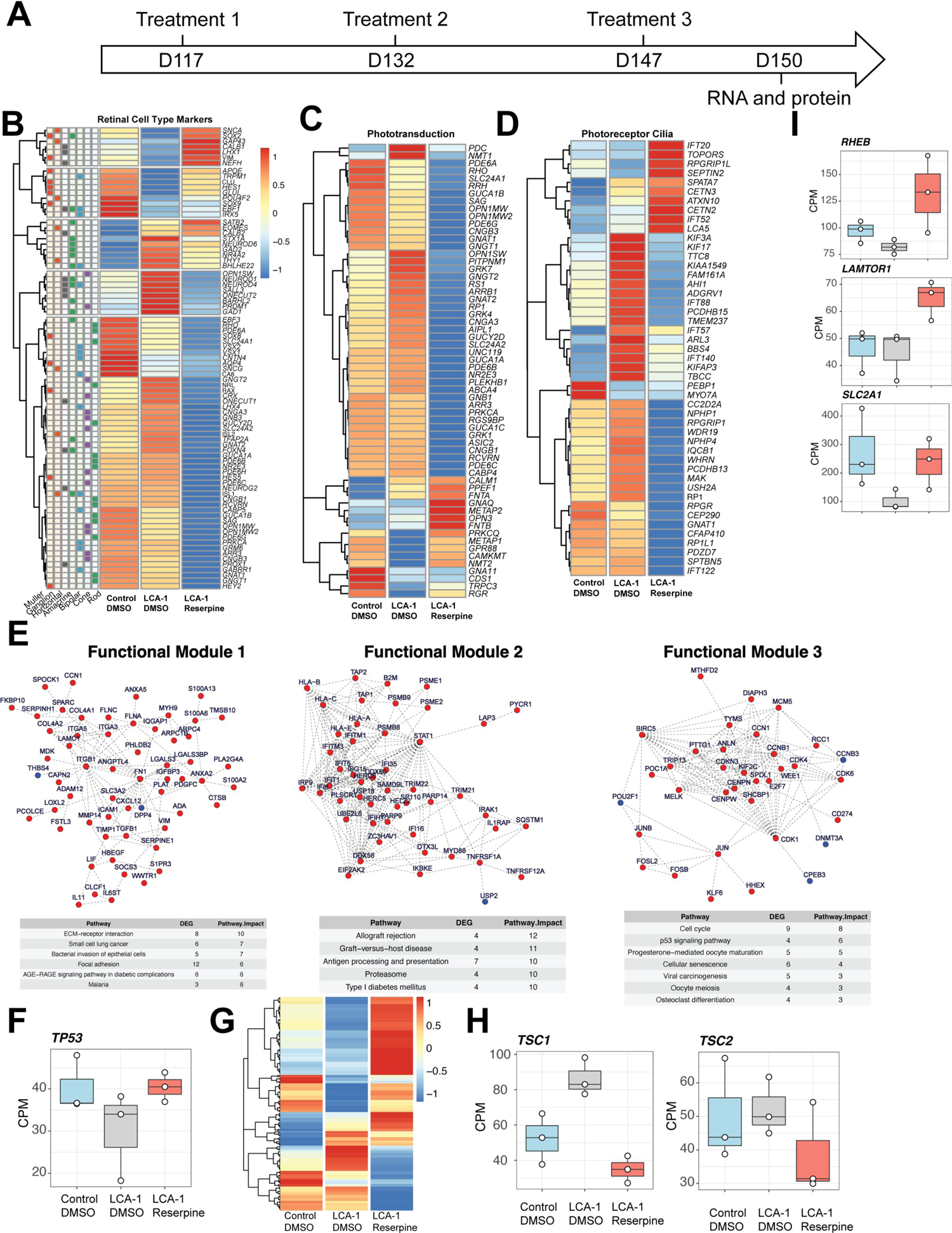
Altered expression of retinal pathway genes by reserpine treatment. (**A**) Timeline of treatment and sample harvest. (**B**) Different retinal cell types appeared to respond uniquely to reserpine. (**C**) Phototransduction genes were globally downregulated except for specific genes like OPN3. (**D**) Genes associated with photoreceptor cilia showed mixed trends. (**E**) Functional clustering of differentially expressed genes show activity in cell survival pathways. (**F**) TP53 gene expression change with reserpine treatment. (**G**) Heatmap of p53 signaling pathway (combination of KEGG, Reactome and Hallmark genes). (**H**) Transcriptional response of TSC1 and TSC2 genes show reduced expression after reserpine treatment. (**I**) Expression of mTOR pathway genes upon reserpine treatment.

**Fig. S6.**
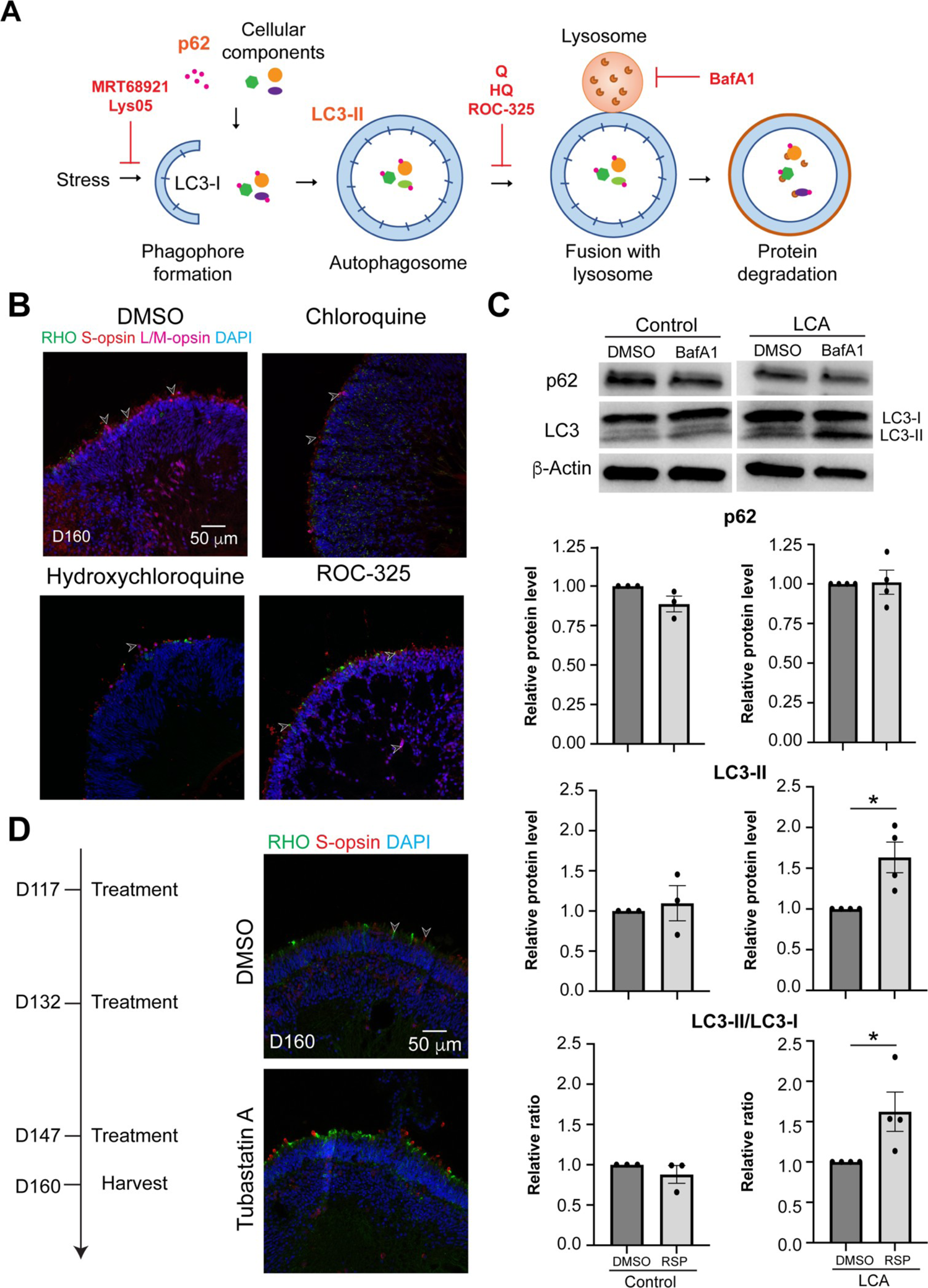
Effect of autophagy modulators on LCA-1 organoids. (A) Schematic diagram of autophagy with annotation of the drug effect of autophagy inhibitors. (B) Immunostaining of rhodopsin (green), S-opsin (red), and L/M-opsin (magenta) was used to evaluate the effect of chloroquine, hydroxychloroquine, and ROC-325. (**C**) Western blot and quantification of p62, LC3-II and LC3-II/LC3-I ratio upon bafilomycin A1 (BafA1) treatment. The drug vehicle dimethylsulfoxide (DMSO) was added to the cultures in the untreated group at the same volume as the drug. b-Actin was used as the loading control. The histograms summarize data in at least 3 batches of experiments, each of which contained at least 3 retinal organoids per group. Each dot in the histogram shows data in one batch of experiment and are presented as mean ± standard error. *, p<0.05. (**D**) Treatment timeline (left) and outcome of Tubastatin A treatment (right) as shown by immunostaining of rhodopsin (green) and S-opsin (red). In all images, nuclei were stained by 4’,6-diamidino-2-phenylindole (DAPI). Arrowheads indicate relevant staining. Images were representative of at least 2 independent experiments, each of which had at least 3 organoids.

**Fig. S7.**
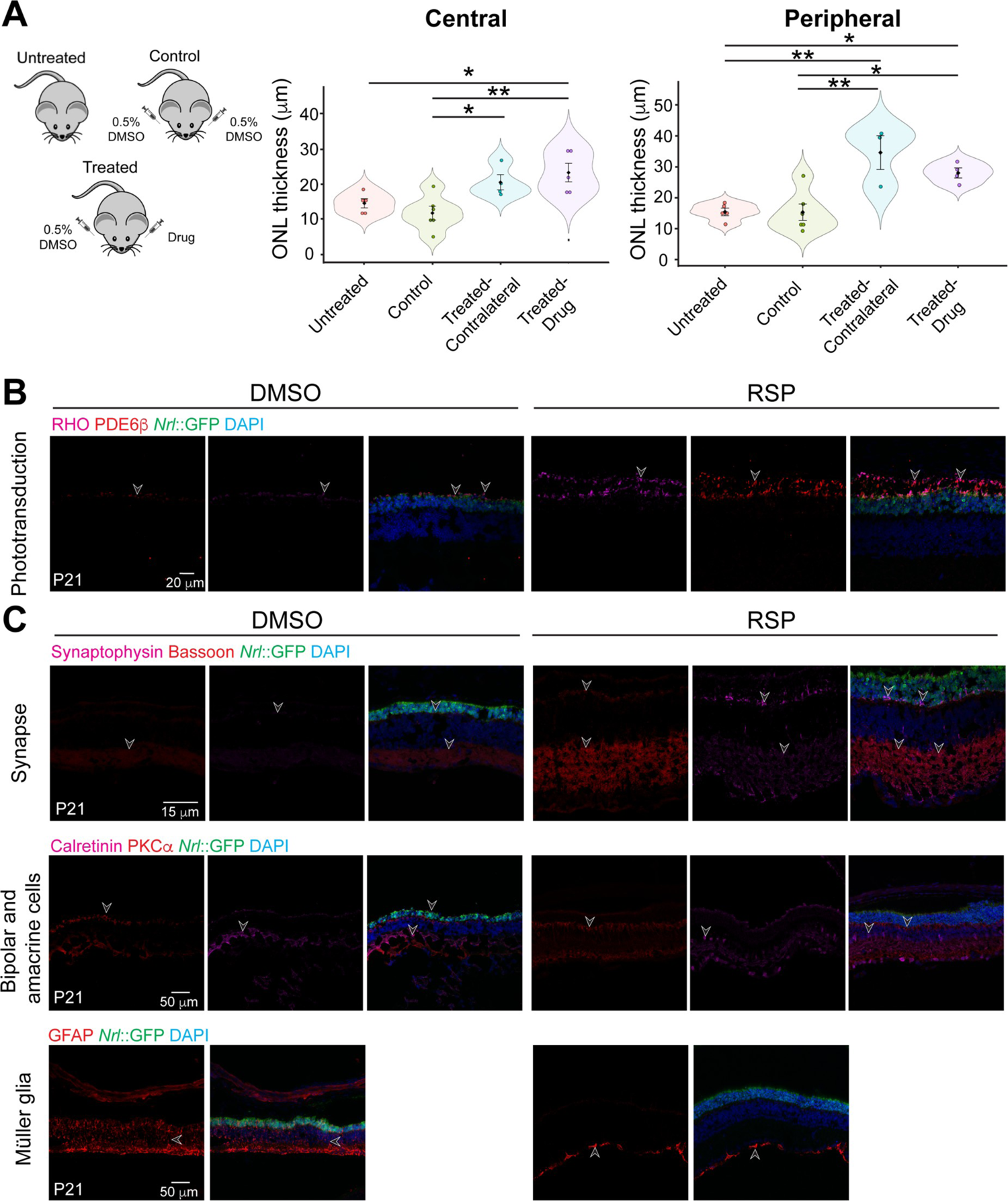
Intravitreal injection of reserpine (RSP) to *rd16* mice. (**A**) Evaluation of the impact of different injection schemes on outer nuclear layer (ONL) thickness of central and peripheral retina. Each dot in the bee swamp plot indicates the ONL thickness of one retina from one mouse. The shape of the plot indicates the distribution of data points, which are shown by colorful circles in the center. The black diamond indicates the mean, and the error bar reveals standard deviation. (**B**) Immunostaining of phototransduction protein rhodopsin (RHO, magenta) and PDE6β (red). (**C**) Retinal structures and cell types shown by synaptophysin for presynaptic vesicles (magenta, top panel), bassoon for ribbon synapses (red, top panel), PKCa for bipolar cells (red, middle panel), Calretinin for amacrine cells (magenta, middle panel), GFAP for cellular stress and Müller glia (red, bottom panel). Nuclei were stained by 4’,6-diamidino-2-phenylindole (DAPI). Arrowheads indicate relevant staining. Images were representative of at least 3 mice, each of which was from a different litter.

**Supplemental Table 1.** Antibody information

| Antibody | Vendor | Catalog # | Host | Dilution |  |
| --- | --- | --- | --- | --- | --- |
|  |  |  |  | IHC* | IB^ |
| Rhodopsin (RHO) | Gift of Dr. Robert Molday, Univ British Columbia |  | Mouse | 1:500 | 1:1000 |
| S-opsin | Santa Cruz | sc-14363 | Goat | 1:500 |  |
| Polyglutamylation Modification (GT335) | AdipoGen | AG-20B-0020 | Mouse | 1:250 |  |
| Rootletin | Chicken | Custom | Chicken | 1:250 |  |
| ADP-ribosylation factor-like protein 13B (Arl13b) | Proteintech | 17711-1-AP | Rabbit | 1:500 |  |
| L/M-opsin (OPN1LMW) | Millipore | AB5405 | Rabbit | 1:500 |  |
| Centrosomal protein 290 (CEP290) | Bethyl laboratories | A301-659A | Rabbit |  | 1:500 |
| Glyceraldehyde-3-phosphate dehydrogenase (GAPDH) | Millipore/Sigma | G8795 | Mouse |  | 1:1000 |

\*IHC, immunohistochemistry
^IB, immunoblotting

